# A molecular surveillance-guided vector control response to concurrent dengue and West Nile virus outbreaks in a COVID-19 hotspot of Florida

**DOI:** 10.1101/2021.10.08.21264776

**Authors:** Heather Coatsworth, Catherine A. Lippi, Chalmers Vasquez, Jasmine B. Ayers, Caroline J. Stephenson, Christy Waits, Mary Florez, André B. B. Wilke, Isik Unlu, Johana Medina, Maria L. Alcaide, Sadie J. Ryan, John A. Lednicky, John C. Beier, William Petrie, Rhoel R. Dinglasan

## Abstract

Simultaneous dengue virus (DENV) and West Nile virus (WNV) outbreaks in Florida, USA, in 2020 resulted in 71 dengue virus serotype 1 and 86 WNV human cases. Our outbreak response leveraged a molecular diagnostic screen of mosquito populations for DENV and WNV in Miami-Dade County to quickly employ targeted mosquito abatement efforts. We detected DENV serotypes 2 and 4 in mosquito pools, highlighting the silent circulation of diverse dengue serotypes in mosquitoes. Additionally, we found WNV-positive mosquito pools in areas with no historical reports of WNV transmission. These findings demonstrate the importance of proactive, strategic arbovirus surveillance in mosquito populations to prevent and control outbreaks, particularly when other illnesses (e.g., COVID-19), which present with similar symptoms are circulating concurrently. Growing evidence for substantial infection prevalence of dengue in competent mosquito vectors in the absence of local index cases suggests a higher level of dengue endemicity in Florida than previously thought.

**Article Summary Line:** Evidence of increasing dengue endemicity in Florida: Vector surveillance during dengue and West Nile virus outbreaks revealed widespread presence of other dengue virus serotypes in the absence of local index cases.

## Introduction

Although West Nile Virus (WNV) is endemic in the continental United States, only a handful of states, especially Florida, are at risk of autochthonous dengue virus (DENV) transmission *(1,2)*. Throughout 2020, Miami-Dade County was the epicenter of COVID-19 in Florida, but unbeknownst to the general U.S. population, the state experienced in parallel, concurrent outbreaks of DENV and WNV from May - December 2020, resulting in 71 local cases of dengue fever (DENV serotype 1) and 86 cases of WNV. Herein, we report the outcome of vector surveillance and control effort in response to the arbovirus outbreaks in Miami-Dade County (August-November 2020).

## Methods

### Sample collection and processing

Adult mosquitoes were collected with BG-Sentinel traps (Biogents AG, Regensburg, Germany) baited with dry ice, for 24 hours. Miami-Dade Mosquito Control District (MDMCD) trapped, morphologically identified, and sorted adult female mosquitoes by location into pools of 2-25 mosquitoes of the same species. Samples were kept cold to avoid RNA degradation, shipped on dry ice, and stored at -80°C. In total, 743 pools were collected for testing [548 for DENV (pools of *Aedes aegypti* and *Aedes albopictus*, N= 5,079 mosquitoes) and 188 for WNV (pools of *Anopheles crucians, Culex coronator, Culex nigripalpus*, and *Culex quinquefasciatus*, N= 2,589 mosquitoes)].

### RNA extraction

Chilled, sterile 1X phosphate-buffered saline (PBS) and sterile glass beads were added to each sample in a biosafety cabinet (BSC). Each 1 mL sample was homogenized in a Bullet Blender (speed 8, 5 minutes) with repeated cooling on ice/cold block in the BSC. Samples were centrifuged (3,750xg, 3 minutes) and 140 µL of the homogenate supernatant was added to AVL lysis buffer (560 µL) (Qiagen). RNA was extracted using the QIAmp Viral RNA extraction kit (Qiagen) following the manufacturer’s protocols with 2×40 µL elution steps. One pool of 5 uninfected *Ae. aegypti* (Orlando) mosquitoes was processed in parallel, serving as a negative extraction control to rule out contamination or spurious amplification.

### Real-Time RT-PCR Virus Detection

Sample RNA was either tested for i) WNV or ii) DENV based on the mosquito species for a given pool. Samples designated for DENV screening were first run through a pan-dengue serotype screen, DENVAll (Appendix Table 1), while the remaining samples were screened via a WNV assay (Appendix Table 2). Each sample, prepared in a PCR hood with static air, was run as technical duplicates. Each plate included a mosquito extraction control, a no template control, and either i) a positive WNV control or ii) dengue virus serotype 1, 2, 3 and 4 (DENV-1 through DENV-4) positive controls. All positive RNA controls were obtained from BEI resources diluted 1:10 (Appendix Tables 1,2,5). Analyses used either QuantaBio UltraPlex 1-Step ToughMix (4X) Low-ROX master mix (Appendix Table 3) or SuperScript™ III Platinum™ One-Step qRT-PCR (Appendix Table 4) on a BioRad CFX96 Touch Real-Time PCR Detection System at 50°C for 30 minutes (for Superscript reactions) or 50°C for 10 minutes (for QuantaBio reactions), 95°C for 2 minutes, and 45 cycles of: 95°C for 15 seconds and 60°C for 45 seconds. Both assays yielded similar results, which helped overcome availability bottlenecks during the COVID-19 pandemic. Samples that were DENV-positive via the DENVAll assay were analyzed for serotype specificity (Appendix Tables 5). Any putative positive or inconclusive (i.e., only one replicate amplified) samples were re-run in confirmatory reactions, adding extra technical replicates and performing additional confirmatory runs as needed to confirm virus positivity.

**Table 1.** Positive mosquito pool results from screening mosquitoes collected in Miami-Dade County (Florida, USA) in 2020. WNV = West Nile virus, DENV-2 = dengue virus serotype 2, DENV-4 = dengue virus serotype 4

| Collection Date | Sample ID | Overall sample result | Species | Pool Size |
| --- | --- | --- | --- | --- |
| 06/09/20 | N/A* | WNV positive | *Culex quinquefasciatus* | 18 |
| 06/23/20 |  | WNV positive | *Culex quinquefasciatus* | 4 |
|  |  | WNV positive | *Culex quinquefasciatus* | 24 |
| 06/30/20 |  | WNV positive | *Culex quinquefasciatus* | 5 |
| 07/23/20 | 0723-30 | WNV positive | *Culex nigripalpus* | 25 |
|  | 0723-31 | WNV positive | *Culex nigripalpus* | 25 |
|  | 0723-39 | WNV positive | *Culex nigripalpus* | 25 |
|  | 0723-44 | WNV positive | *Culex nigripalpus* | 25 |
| 07/30/20 | 0730-37 | DENV-2 positive | *Aedes aegypti* | 25 |
|  | 0730-41 | DENV-2 positive | *Aedes aegypti* | 25 |
| 08/20/20 | 0820-06 | WNV positive | *Culex nigripalpus* | 25 |
|  | 0820-07 | WNV positive | *Culex nigripalpus* | 25 |
|  | 0820-08 | WNV positive | *Culex nigripalpus* | 25 |
|  | 0820-09 | WNV positive | *Culex nigripalpus* | 25 |
|  | 0820-11 | WNV positive | *Culex nigripalpus* | 25 |
| 08/28/20 | 0828-2 | DENV-2 positive | *Aedes aegypti* | 23 |
|  | 0828-10 | DENV-2 positive | *Aedes aegypti* | 25 |
| 09/17/20 | 0917-4 | DENV-4 positive | *Aedes aegypti* | 13 |
|  | 0917-8 | DENV-4 positive | *Aedes aegypti* | 8 |
|  | 0917-11 | DENV-4 positive | *Aedes aegypti* | 8 |
|  | 0917-12 | DENV-4 positive | *Aedes aegypti* | 4 |
|  | 0917-13 | DENV-4 positive | *Aedes aegypti* | 10 |
|  | 0917-16 | DENV-4 positive | *Aedes aegypti* | 3 |
|  | 0917-27 | DENV-4 positive | *Aedes aegypti* | 6 |
| 09/22/20 | 0922-2 | DENV-4 positive | *Aedes aegypti* | 5 |
| 09/29/20 | 0929-07 | DENV-4 positive | *Aedes aegypti* | 2 |

\*Pathogen detection conducted by the CDC.

### Positive and negative sample designation

A sample was considered ‘positive’ based on positive technical duplicate results in two independent runs using the WNV assay (Appendix Table 2) and the DENV serotype-specific assay (Appendix Table 5). Samples were considered ‘negative’ based on no detectable Ct value in either technical duplicate after 45 cycles.

### Mapping mosquito samples

To understand the spatial distribution of mosquito pools collected for this effort, we developed maps using ArcMap (v 10.6) at the zip code level, which is the smallest level of resolution obtainable with our datasets. All datasets were either publicly available: Miami-Dade County Boundaries, population density and land use (2010 US Census, Miami-Dade County’s Open Data Hub), median household income (2010 US Census, Michigan Population Studies), persons living with HIV/AIDS (PLWH) (2018, AIDsVu), or obtained with permission from personal communications (2018 – 2020 mosquito prevalence c/o Chalmers Vasquez, Miami-Dade Mosquito Control District; 2009 – 2019 Imported DENV cases c/o Andrea Morrison, Florida Department of Health). Land-use types were manually concatenated to 11 different primary categories: Agricultural, Cemeteries, Commercial, Educational, Industrial, Marine, Recreational, Residential, Paved, Water, and Vacant. These types were further summed as: ‘urban/built’ (Commercial, Educational, Industrial, and Paved), ‘agricultural/recreational’ (Agricultural, Recreational), ‘residential’ (Residential) and ‘other’ (Cemeteries, Marine, Water, and Vacant) (Appendix Table 6).

### Spatial statistical analysis of vector distribution and spatial visualization of overlapping PLWH and arbovirus hotspots

To explore the spatial relationship between positivity and overall mosquito sampling, the directional distribution and mean center (e.g., central tendencies, dispersion) were computed for DENV-and WNV-positive mosquito pools, for all collected DENV vectors (*Ae. aegypti* and *Ae. albopictus*) and all WNV vectors (*An. crucians, Cx. coronator, Cx. nigripalpus*, and *Cx. quinquefasciatus*), both individually and together. Kernel density estimation (KDE) with optimal distance bandwidths *(3)* was used to visualize the continuous density of DENV- and WNV-positive mosquito pools, as well as traps containing un-infected DENV and WNV vectors. To determine whether arbovirus-positive pools occurred in areas where there were imported DENV cases or in areas with a higher number of PLWH, local Moran’s I, a local indicator of spatial association (LISA), analyses with inverse distance weighting were performed to detect and identify PLWH hotspots (areas of elevated incidence or prevalence) that potentially overlap with locations of arbovirus positive pools, as well as imported DENV cases for 2019 only, and for the 2009-2019 period. All spatial statistical analyses were completed in ArcMap (v 10.6).

## Results

### Molecular detection of arbovirus-positive mosquito pools

We found that 2.96% of female mosquito pool samples (22/743 pools, 382/7,668 mosquitoes) were positive for an arbovirus; 4 were DENV-2 positive, 9 were DENV-4 positive (2.43% DENV positivity), and 9 were WNV positive (4.79% WNV positivity) (Table 1 and Appendix 7). Results were confirmed and verified independently by the Florida Department of Health (FLDOH), and the inclusion of a pool of lab reared mosquitoes in every extraction and PCR run ruled out contamination during lab handling or spurious amplification of mosquito material.

For DENV, we tested 45 *Ae. albopictus* and 510 *Ae. aegypti* mosquito pools, and only *Ae. aegypti* pools were found to be positive for DENV-2 and/or DENV-4 (2.43% positivity).

For WNV, we tested five *An. crucians*, five *Cx. coronator*, 47 *Cx. nigripalpus* and 131 *Cx. quinquefasciatus* pools. Only *Cx. nigripalpus* pools were WNV positive (19.1% positivity), although CDC findings from earlier in 2020 (June), found three positive *Cx. quinquefasciatus* pools.

Mosquito pool samples that were found to be DENV- or WNV-positive were reported to the MDMCD within 48-72 hours. Miami-Dade Mosquito Control Division initiated door-to-door source reduction in the areas where positive mosquito pools were found. Vectobac WGD (Valent Biosciences), a larvicide product containing *Bacillus thuringiensis israelensis*, was applied at positive pool sites for four weeks. Adulticide treatment using chlorpyrifos was also conducted (weather permitting) using Mosquitomist (Clarke).

### A spatial-climatic distribution of arbovirus-positive mosquito pools identified exigent and emerging areas of concern

DENV-positive mosquito pools were distributed throughout the county, with most (11/13) positive pools located in downtown Miami (N=3), Wynwood (N=2), and South Miami (N=6) (Figure 1A) and in areas with mid-to-high human population density (2,000 – 7,000 people/km^2^) (Figure 1B). At the zip code level, DENV-positive pools were primarily found in built and residential environments (59.42%) over more rural, (i.e., agricultural and recreational) areas (14.62%) (Figure 1C). Conversely, the thirteen WNV-positive pools (9 identified herein) had three primary infection foci – in Wynwood (N=4), high population density (>3000 people/km^2^, urban built environment), in Homestead (N=8), low population density (<470 people/km^2^, mostly agricultural and recreational land), and in North Miami (N=1), mid population density (1,000 – 3,000 people/km^2^ and mixed land use) (Figure 1D-F). WNV-positive pools were primarily from agricultural and recreational areas (69.32%) and were less common in urban built and residential areas (15.98%).

**Figure 1.**
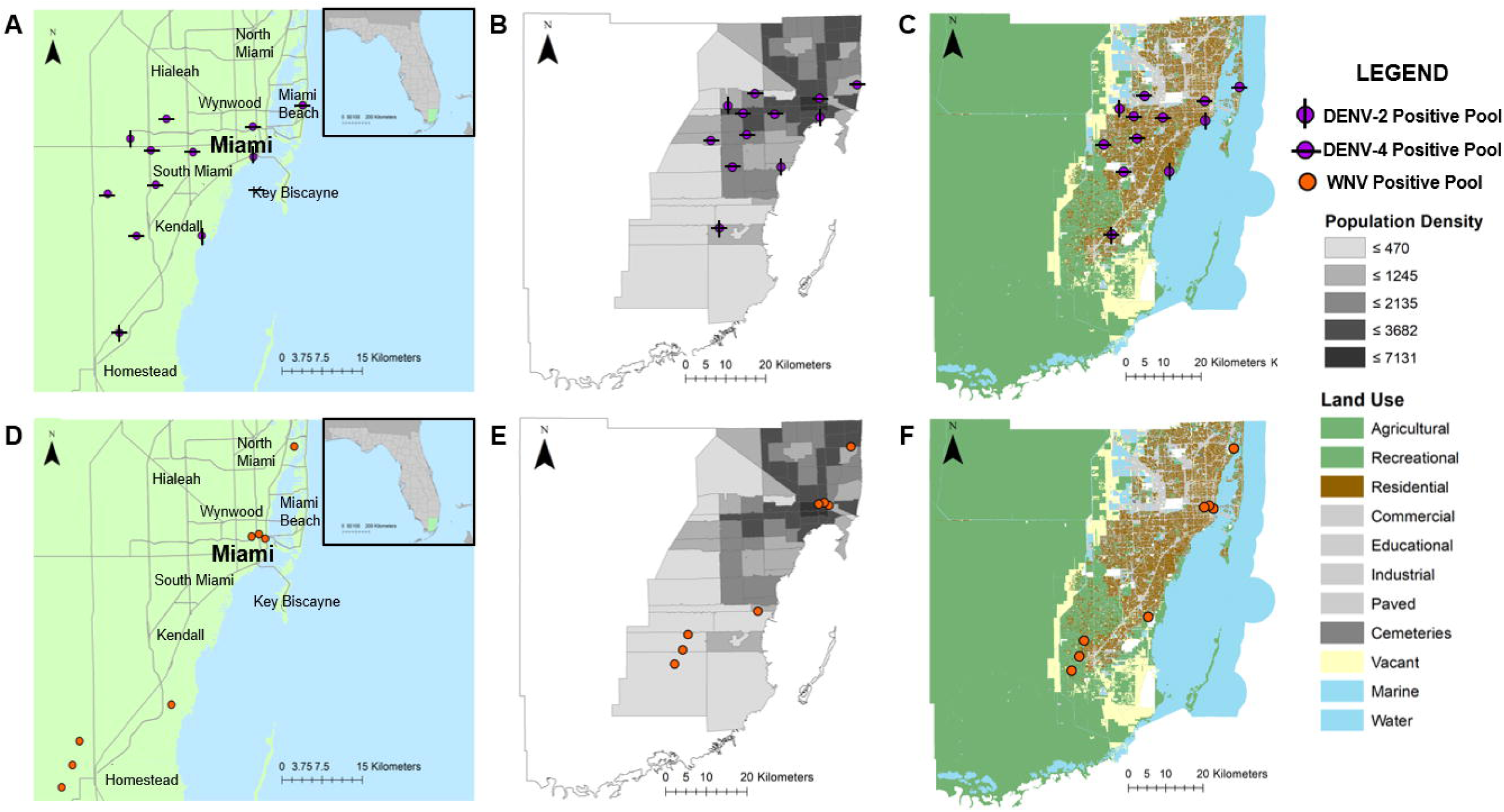
Spatial distribution of arbovirus positive mosquito pools (purple – DENV, orange – WNV). A and D: Positive pool spread throughout Miami-Dade County – DENV positive pools (A), or WNV positive pools (D). B and E: Positive pool spread overlayed on 2010 Miami-Dade population density (individuals/km2) – DENV positive pools (B), or WNV positive pools (E). C and F: Positive pool spread overlayed on 2010 Miami-Dade land use data – DENV positive pools (C) or WNV positive pools (F).

The directional distribution and mean center computations of dengue vector distributions showed that DENV-positive mosquito pools were within the directional distribution of all dengue vector traps (Figure 2A). *Ae. aegypti* prevalence dominated the traps in much of the county and had a much larger directional distribution than *Ae. albopictus*, whose presence was concentrated in southern Miami-Dade County (Figure 2B). Although the directional distribution of all WNV vectors overlapped with the directional distribution of the WNV-positive mosquito pools, the latter extended well beyond the southernmost tip of the WNV vector directional ellipse (Figure 2C). *Culex quinquefasciatus, An. crucians* and *Cx. nigripalpus* had distinct directional distributions in the Miami Beach area, North Miami, and South Miami, respectively (Figure 2D). The distribution of *Cx. coronator* encompassed all three of these zones, lacking any strict boundary (Figure 2D).

**Figure 2.**
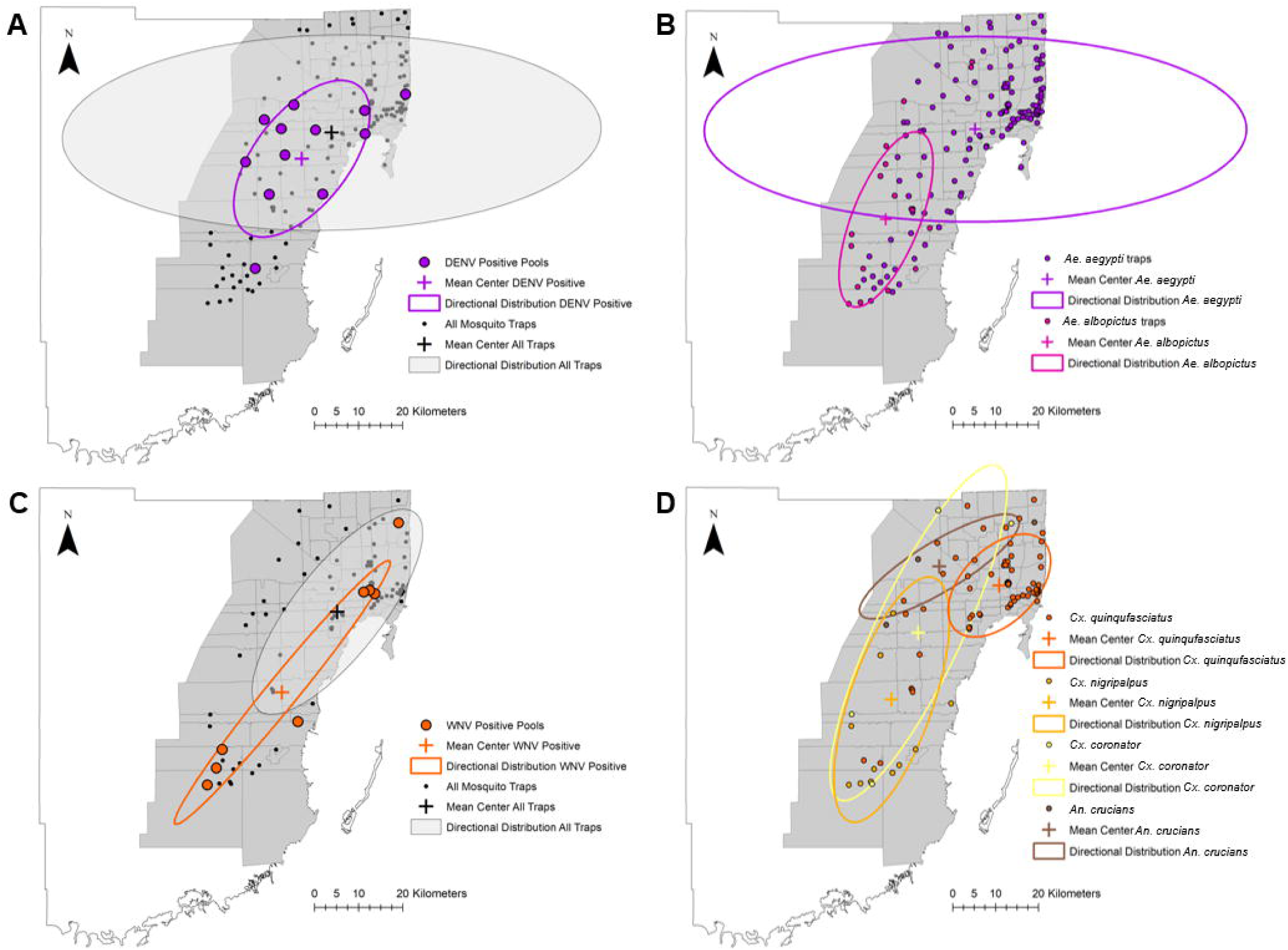
Spatial analysis of A: DENV positive pool distribution (purple) within traps containing DENV vectors (black), B: Distribution of traps containing *Aedes aegypti* (purple) or *Aedes albopictus* (pink), C: WNV positive pool distribution (orange) within traps containing WNV vectors (black), D: Distribution of traps containing *Culex quinquefasciatus* (dark orange), *Culex nigripalpus* (light orange), *Culex coronator* (yellow), or *Anopheles crucians* (brown).

The KDE maps show a clear high density of vector trapping efforts across Miami-Dade County except for the southernmost area (Appendix Figure 1A). The KDE map for DENV-2 positive pools showed high values surrounding each positive pool but low values between pools (Appendix Figure 1B), while the DENV-4 positive KDE map showed connected high values across all positive pools (Appendix Figure 1C). The KDE map for the WNV vector trapping effort showed a high concentration of vectors in the downtown Miami/Miami Beach areas, with areas of mid values dotted throughout the county (Appendix Figure 1E). The WNV-positive pools showed two clear high-value foci, one in Homestead, and one in Miami Beach (Appendix Figure 1F).

DENV-positive mosquito pools were in areas of higher average maximum temperatures, lower average minimum temperatures, and higher total precipitation (Appendix Figure 2 A-C). WNV-positive pools displayed the same trend with respect to temperature but were found in areas of lower total precipitation (Appendix Figure 2D-F).

LISA analyses on imported DENV case data showed similar clusters for both the 2009-2019 imported DENV data (Appendix Figure 3A), as well as the 2019 only imported DENV dataset (Appendix Figure 3B). The northwestern area of Miami-Dade was a high-high cluster, indicating this area is a hotspot for imported DENV cases. Conversely, the Miami Beach area was a low-low cluster, suggesting a consistently lower-than-average number of imported DENV cases (a cold spot). The low-high outliers for both datasets occurred just below the hotspot in northwest Miami-Dade, while the high-low outlier for the decade-wide dataset was in South Miami, and the 2019 only data had high-low outliers throughout North Miami.

We explored the possibility of increased arbovirus risk to special populations in Miami-Dade (i.e., PLWH) by examining in parallel the LISA results for PLWH and the spatial distribution of DENV or WNV positive pools and observed that DENV-positive mosquito pools were in the low-low PLWH cluster (Appendix Figure 3C), while one of the WNV-positive pool foci was in a PLWH hotspot (Appendix Figure 3D).

## Discussion

### Ecological distribution of WNV-positive mosquito pools

Although WNV-positive mosquito pools are rarely reported in Miami-Dade County, we found nine positive mosquito pools collected between June – November 2020; three other WNV positive mosquito pools were found in 2020 (collected earlier in June and analyzed by the CDC). All four mosquito species we tested can transmit WNV *(4,5)*; however, we only found WNV-positive *Cx. nigripalpus* pools, an expected result, as it is the primary enzootic and epidemic vector of mosquito-borne viruses encephalitides such as WNV, St. Louis encephalitis virus, and eastern equine encephalitis virus throughout southern Florida *(6)*. This result is intriguing, considering the overwhelmingly dominant collection of *Cx. quinquefasciatus* throughout the county. Differences in WNV vector positivity may be due to several factors including host preference and vector competence. Previous reports show that *Cx. quinquefasciatus* has a wide range of WNV competency that appears to partition according to spatial and climatic scales and is influenced by virus genetic background *7–11)*.

There appeared to be a centralized distribution of traps collecting *Culex* and *Anopheles* mosquitoes, while positive pool traps were dispersed throughout the county. The spatial partitioning of the WNV-positive mosquito pools suggests that the positive pools had a distinct spatial trend within the larger distribution of traps collecting *Culex* and *Anopheles* mosquitoes in Miami-Dade County. This might indicate that current trap coverage is not adequate to reliably detect WNV-positive mosquito pools. As WNV infects a wealth of other non-human animals (i.e., horses, alligators, birds, etc.) *(12)*, controlling the spread of WNV requires additional zoonotic measures including implementing equine WNV vaccines, as well as testing dead birds to understand prevalence and virus hotspots throughout the county.

WNV-positive mosquito pools were primarily located in areas with lower precipitation, higher maximum temperature, and lower minimum temperature (range of 21-29°C), optimal for multiple *Culex* vectors *(10)*. As there was no clear overlap between the spatial distribution of WNV vectors and climatic variables, these general climatic conditions are likely ideal for all the WNV vectors we analyzed. WNV-positive mosquito pools were found primarily in agricultural and recreational areas. These areas include protected bird sanctuaries, as well as habitats for resident and migrating shorebirds, which are known WNV avian hosts *(12)*.

### Ecological distribution of DENV-positive mosquito pools

Although other dengue vectors (i.e., *Aedes spp*.) are present in Miami-Dade *(13)*, our data suggest that *Ae. aegypti* is clearly the primary vector of concern. Evidence suggests that Floridian *Ae. aegypti* (Monroe County) and *Ae. albopictus* (Indian River County) are similarly competent for DENV-1 *(14)*. DENV vector competency varies greatly based on DENV serotype and FL *Ae. aegypti* geographic origin *(15)*, so extrapolations of *Ae. aegypti* and *Ae. albopictus* vector competence across FL may not be appropriate. Our findings may be a by-product of *Ae. aegypti* prevalence, as considerably more *Ae. aegypti* were collected than *Ae. albopictus* in traps across the entirety of Miami-Dade County in 2020, and in former years *(13)*.

DENV-positive pools were primarily located in areas with higher precipitation, higher maximum temperature, and lower minimum temperature. The directional distribution of DENV-positive pools suggests that the current trap spread is more than adequate to reliably detect DENV-positive pools. However, since trap density within the DENV-positive pool distribution area is not as concentrated as in the other *Aedes* positive trap locales, additional traps in the ellipse margins could prove useful.

### Epidemiological relevance of WNV-positive mosquito pools

WNV-positive mosquito pools overlapped with the timing of 27 human symptomatic cases (June, July, August), 33 human asymptomatic blood donors (June, July August), as well as 15 WNV-positive birds (June, July), all in Miami-Dade County *(16)*. One of the WNV-positive pool foci overlapped with a hotspot of PLWH (Appendix Figure 3D). This is especially concerning for PLWH, as these individuals are at a higher risk of neurotropism from WNV infection *17–21)*. WNV positive pools were primarily found in more rural areas with lower median household incomes, and low to medium population density (Figure 1 D-F). Low-income areas have previously been associated with higher WNV prevalence *(22,23)*. These data underscore the need for tailored programs to protect and prevent infection in these at-risk populations.

### Epidemiological relevance of DENV-positive mosquito pools

In the last five years, the number of local DENV cases in Florida has risen from zero (2017) to 71 (2020). A rise in cases can also be seen in the imported DENV case numbers (DENV-1 through DENV-4). In 2017, there were only 18 imported cases, 73 in 2018, 395 in 2019, 41 in 2020. This spike in dengue cases was mirrored in Miami-Dade County, where local case numbers moved from zero cases in 2017, to one in 2018, fourteen in 2019 and four in 2020, and travel-associated cases progressed from nine in 2017, to thirty-eight in 2018, 226 in 2019 and twenty in 2020. The large spike in imported cases in 2019 may have introduced other DENV serotypes into the resident Miami-Dade mosquito populations, acting as seeding events, which are known drivers of local DENV case incidence *(24,25)*.

To examine whether this could be the case, we analyzed the available zip code-mapped historical (2009 – 2019) and 2019 imported DENV data using LISA analyses (Appendix Figure 3A and B), as well as the 2019 only imported DENV dataset (Appendix Figure 3B). The similarity in the decade-long and 2019 LISA analyses suggests that the high number of imported cases in 2019 may have driven the hotspot trends seen in our analyses. None of our DENV-positive mosquito pools overlapped with known imported DENV case hotspots. This could be due to a lack of reporting of zip code level imported DENV data. Alternatively, if mosquitoes bite individuals outside of their listed area of primary residence, such as where individuals spend time outdoors, where they work, or where they socialize, the direct overlap would not be observed.

Local human cases of DENV-1, DENV-2 and DENV-3 have been reported in FL *(2,16,26–28)*. In 2020, only DENV-1 local cases were reported, despite our findings of DENV-2 and DENV-4 positive mosquito pools in the county. ‘Silent’ DENV circulation, defined as transmission between DENV asymptomatic individuals, as well as DENV maintenance in the vector population despite no reported human infection, likely represents the majority of transmission events *(29)*. Finding additional DENV serotypes in the absence of a local human index case is not the first instance of silent DENV circulation in *Ae. aegypti* in the Americas. Previous reports have shown that DENV-4 was found in Manatee County, FL in 2018, and DENV-3 was detected in *Ae. aegypti* in Brazil despite no human index cases *(30,31)*. This silent circulation is likely due to low but persistent vertical transmission in the mosquito population *31–33)*. This disparity could also be due to inherent differences in mosquito vector competence for DENV-1, -2 and -4, as mosquitoes collected in Miami-Dade had higher DENV-1 infection and transmission rates than mosquitoes from Miami-Dade infected with DENV-2 and -4 *(15)*. Although the pathogenicity of the 2020 Miami-Dade human DENV-1 index strain is unknown, the strain may be more infectious to or cause increased disease severity in humans than other circulating dengue strains.

Having multiple concurrent circulating serotypes puts individuals at an increased risk of DENV infection due to complications arising from immune enhancement *(34,35)*. These risks include dengue fever and severe dengue (dengue hemorrhagic fever and dengue shock syndrome) and can be fatal *(36)*. A woman in her 30’s died from DENV-2 in Miami-Dade in 2019, and subsequent viral analysis suggested that infection occurred after DENV circulation had occurred in the Miami area *(37)*.

There were more DENV-positive pools in areas with lower prevalence of PLWH, higher median household income and mid-high population density (Figure 1A-C, Appendix Figure 3C). Although there does not seem to be evidence for an association between income and DENV prevalence *(38)*, DENV is a known urban-centric disease *(39)*.

### The perfect storm: arbovirus transmission in a global pandemic

Miami Dade County has been a hotspot of SARS-CoV-2 transmission throughout the pandemic, with the highest case rate and death toll in the state. Unfortunately, due to the non-standardized nature of COVID-19 case reporting throughout Florida (reports range from residence-based, testing locale-based and exposure-based), we could not directly map COVID-19 prevalence alongside mosquito pool positivity. The potential for immunocompromise resulting from acute SARS-CoV-2 infection to increase severity and/or transmissibility of other diseases (i.e. West Nile, dengue) in the Miami Dade population is concerning.

Due to the similar symptoms produced by COVID-19, West Nile, and dengue infection (fever, headache, muscle pain, nausea, vomiting, malaise), symptomatic WNV or DENV infected individuals may have self-quarantined assuming they were positive for COVID-19 and never received a correct diagnosis. The high number of asymptomatic WNV positive individuals identified directly through blood donorship suggests that the proportion of asymptomatic WNV positive human carriers may be high, and previous reports show that the majority of DENV infected humans are also asymptomatic carriers *(29)*. It should be noted that dengue is not currently part of routine blood donor screening in Miami-Dade.

The spread of the positive pools is intriguing, as prior arbovirus positive pools (for WNV and Zika virus) in Miami-Dade County were solely concentrated within the Miami Beach, Little River and Wynwood locales. Our data suggest a much wider arbovirus positive pool spread, ranging across the county.

Together, these data highlight a lack of vector containment and the importance of continued surveillance for arboviruses in their vector species. This is especially true when infection risk and continued transmission is increased due to: i) the presence of other illnesses (e.g., COVID-19) that present similarly in a clinic to arbovirus infection, ii) the high incidence of PLWH in areas of known WNV positivity resulting in potentially worse health outcomes following infection, and iii) climatic conditions favoring above-average *Ae. aegypti* and *Cx. nigripalpus* vector indices. The large proportion of asymptomatic WNV cases highlights the critical need for improved DENV diagnostics, as no dengue rapid diagnostic tests are currently cleared for use in the United States *(37)*. Prevention and control measures are imperative to prevent future and expanded DENV and WNV outbreaks; targeted vector control efforts through continued mosquito pathogen screening and subsequent population specific insecticide spraying are necessary to prevent arbovirus spread throughout the county.

## Data Availability

All data produced in the present work are contained in the manuscript.

## Acknowledgments

This research was supported in part by U.S. Centers for Disease Control and Prevention (CDC) grant 1U01CK000510-03, Southeastern Regional Center of Excellence in Vector Borne Diseases Gateway Program. The CDC did not have a role in the design of the study, the collection, analysis, or interpretation of data, or writing the manuscript. Christy Waits is a contractor employee of the U.S. Government. This work was prepared as part of her official duties. Title 17, U.S.C., §105 provides that copyright protection under this title is not available for any work of the U.S. Government. Title 17, U.S.C., §101 defines a U.S. Government work as a work prepared by a military Service member or employee of the U.S. Government as part of that person’s official duties. The views expressed in this article are those of the authors and do not necessarily reflect the official policy or position of the Department of the Navy, Department of Defense, nor the U. S. Government. The Department of the Navy, Department of Defense, or the U. S. Government did not have a role in the design of the study, the collection, analysis, or interpretation of data, or writing of the manuscript.

## Author Bio

Dr. Coatsworth is a post-doctoral associate at the University of Florida. Her primary research interests include arbovirology, vector biology, and vector-borne disease transmission.

## Supplementary Information

### Contamination troubleshooting

We noted issues with DENV-2 amplification in the no template and mosquito extraction negative controls with the CDC DENV-2 serotype-specific RT-qPCR assay for a few of the mosquito pools. We addressed these concerns using standard contamination troubleshooting protocols: preparing all qPCR reactions in a PCR hood, deep cleaning qPCR preparation areas, creating new primer and probe dilutions, and using different primer and probe stocks. We also tested each assay reagent for inherent DENV-2 RNA/cDNA contamination but did not find any evidence of contamination. Due to these issues, a new DENV-2 serotype specific assay was implemented (see DENV2 Alm, Supplementary Table 5) targeting a different region of the DENV-2 E glycoprotein. No amplification of negative controls was seen with the new primer-probe combination.

## Appendix

For Appendix Tables 1, 2. and 5: Positive controls used were the following: DENV-1:NR-50530, DENV-2: NR-50531, DENV-3: NR-50532, DENV-4: NR-50533, and WNV: NR-50434. Limits of quantification (LOQs) were determined by standard curve analysis of diluted RNA positive controls

**Appendix Table 1.**
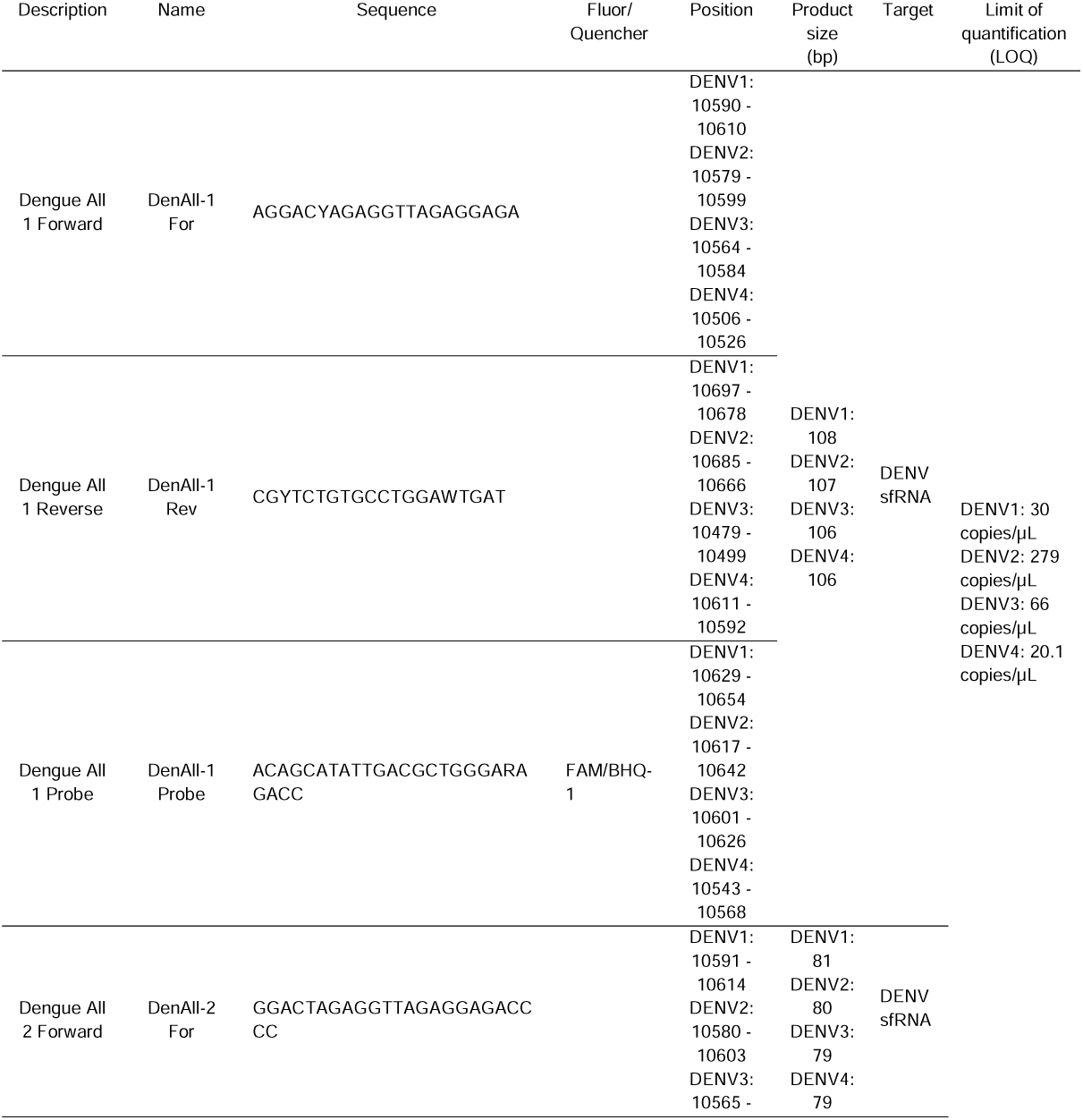
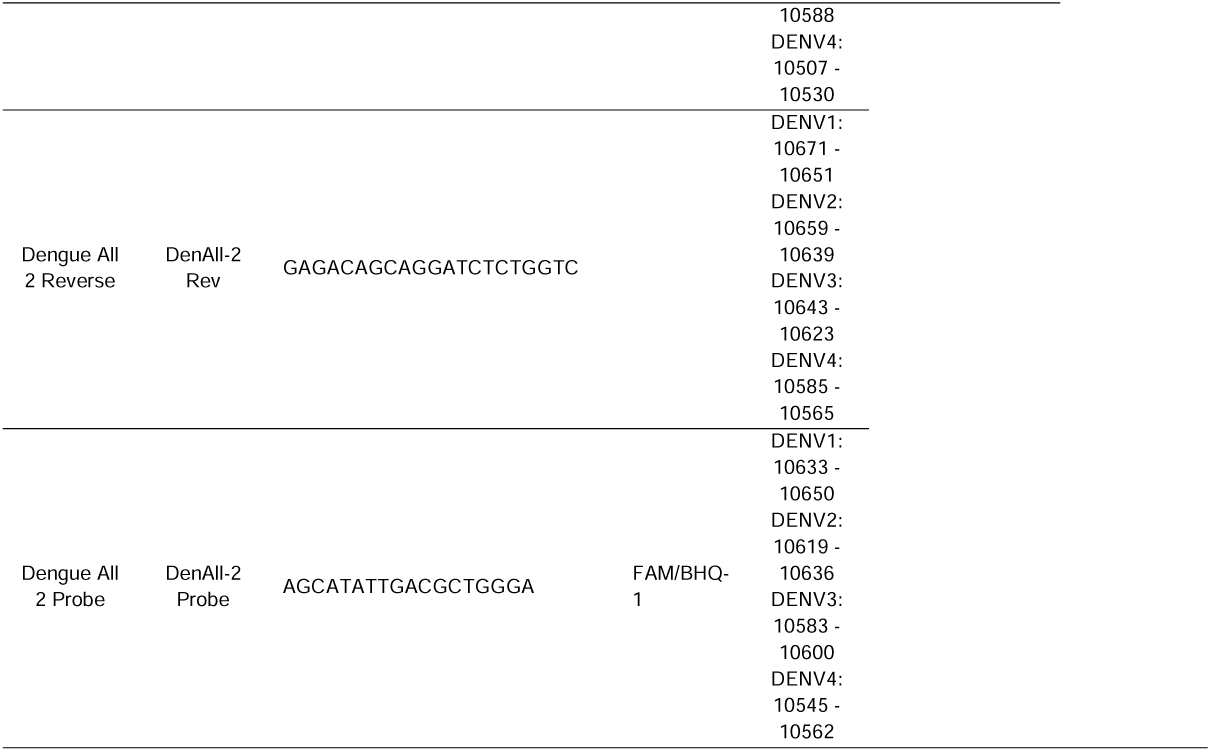
Pan-dengue Real-Time RT-PCR assay (assay courtesy of an active collaboration with Dr. Remi Charrel (Aix Marseille University)

**Appendix Table 2.**
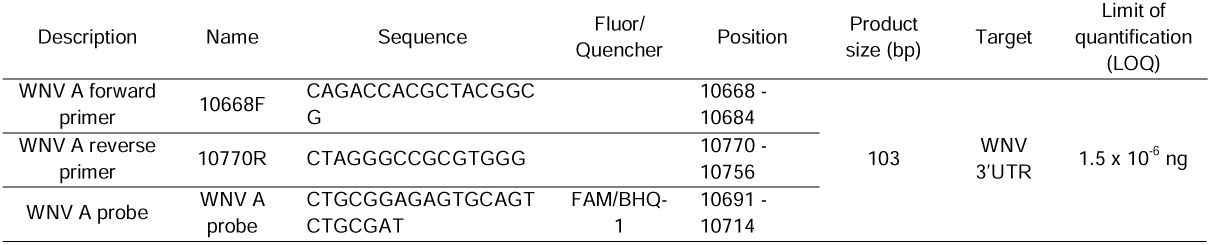
West Nile Virus Real-Time RT-PCR assay (courtesy of Dr. Thomas Unnasch, University of South Florida)

**Appendix Table 3.**
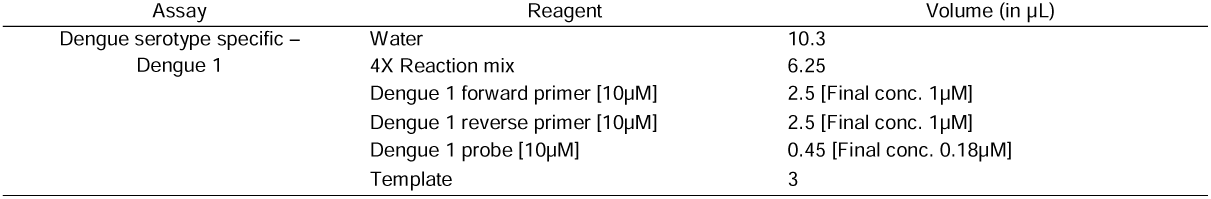
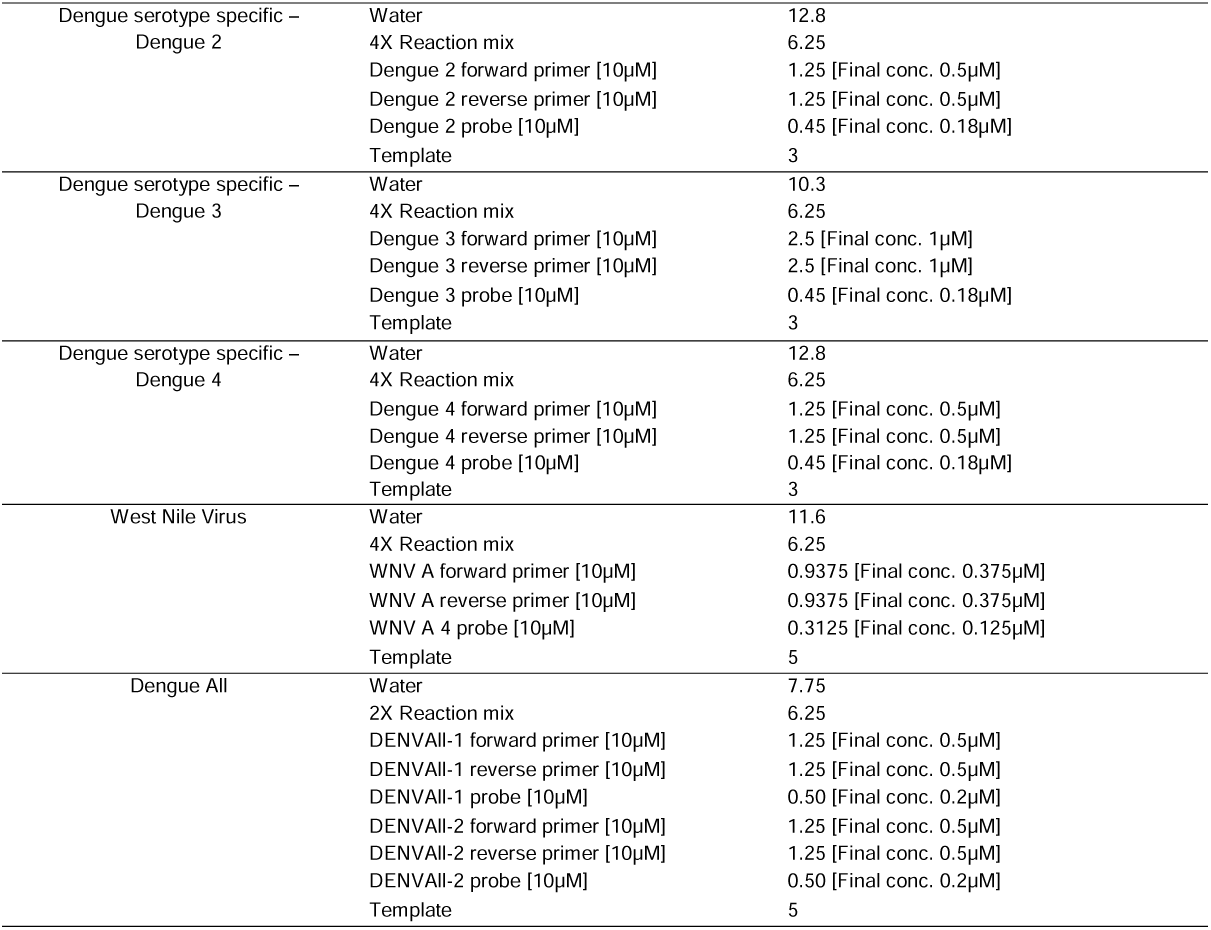
Reaction volumes for each assay using 1-step qRT-PCR using QuantaBio UltraPlex 1-Step ToughMix (4X) Low-ROX master mix. Volumes are representative of one 25µL reaction. Primer and probes were diluted to working solutions of 10µM for assay use. Final primer and probe concentrations/reaction are denoted by [ ]

| Assay | Reagent | Volume (in µL) |
| --- | --- | --- |
| Dengue serotype specific –<br>Dengue 1 | Water | 10.3 |
|  | 4X Reaction mix | 6.25 |
|  | Dengue 1 forward primer [10µM] | 2.5 [Final conc. 1µM] |
|  | Dengue 1 reverse primer [10µM] | 2.5 [Final conc. 1µM] |
|  | Dengue 1 probe [10µM] | 0.45 [Final conc. 0.18µM] |
|  | Template | 3 |
| Dengue serotype specific –<br>Dengue 2 | Water | 12.8 |
|  | 4X Reaction mix | 6.25 |
|  | Dengue 2 forward primer [10µM] | 1.25 [Final conc. 0.5µM] |
|  | Dengue 2 reverse primer [10µM] | 1.25 [Final conc. 0.5µM] |
|  | Dengue 2 probe [10µM] | 0.45 [Final conc. 0.18µM] |
|  | Template | 3 |
| Dengue serotype specific –<br>Dengue 3 | Water | 10.3 |
|  | 4X Reaction mix | 6.25 |
|  | Dengue 3 forward primer [10µM] | 2.5 [Final conc. 1µM] |
|  | Dengue 3 reverse primer [10µM] | 2.5 [Final conc. 1µM] |
|  | Dengue 3 probe [10µM] | 0.45 [Final conc. 0.18µM] |
|  | Template | 3 |
| Dengue serotype specific –<br>Dengue 4 | Water | 12.8 |
|  | 4X Reaction mix | 6.25 |
|  | Dengue 4 forward primer [10µM] | 1.25 [Final conc. 0.5µM] |
|  | Dengue 4 reverse primer [10µM] | 1.25 [Final conc. 0.5µM] |
|  | Dengue 4 probe [10µM] | 0.45 [Final conc. 0.18µM] |
|  | Template | 3 |
| West Nile Virus | Water | 11.6 |
|  | 4X Reaction mix | 6.25 |
|  | WNV A forward primer [10µM] | 0.9375 [Final conc. 0.375µM] |
|  | WNV A reverse primer [10µM] | 0.9375 [Final conc. 0.375µM] |
|  | WNV A 4 probe [10µM] | 0.3125 [Final conc. 0.125µM] |
|  | Template | 5 |
| Dengue All | Water | 7.75 |
|  | 2X Reaction mix | 6.25 |
|  | DENVAll-1 forward primer [10µM] | 1.25 [Final conc. 0.5µM] |
|  | DENVAll-1 reverse primer [10µM] | 1.25 [Final conc. 0.5µM] |
|  | DENVAll-1 probe [10µM] | 0.50 [Final conc. 0.2µM] |
|  | DENVAll-2 forward primer [10µM] | 1.25 [Final conc. 0.5µM] |
|  | DENVAll-2 reverse primer [10µM] | 1.25 [Final conc. 0.5µM] |
|  | DENVAll-2 probe [10µM] | 0.50 [Final conc. 0.2µM] |
|  | Template | 5 |

**Appendix Table 4.**
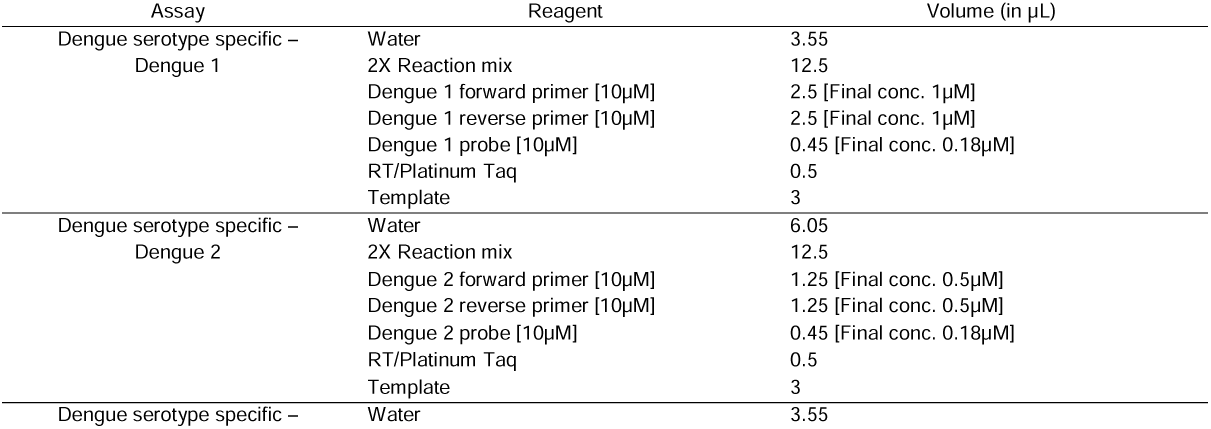
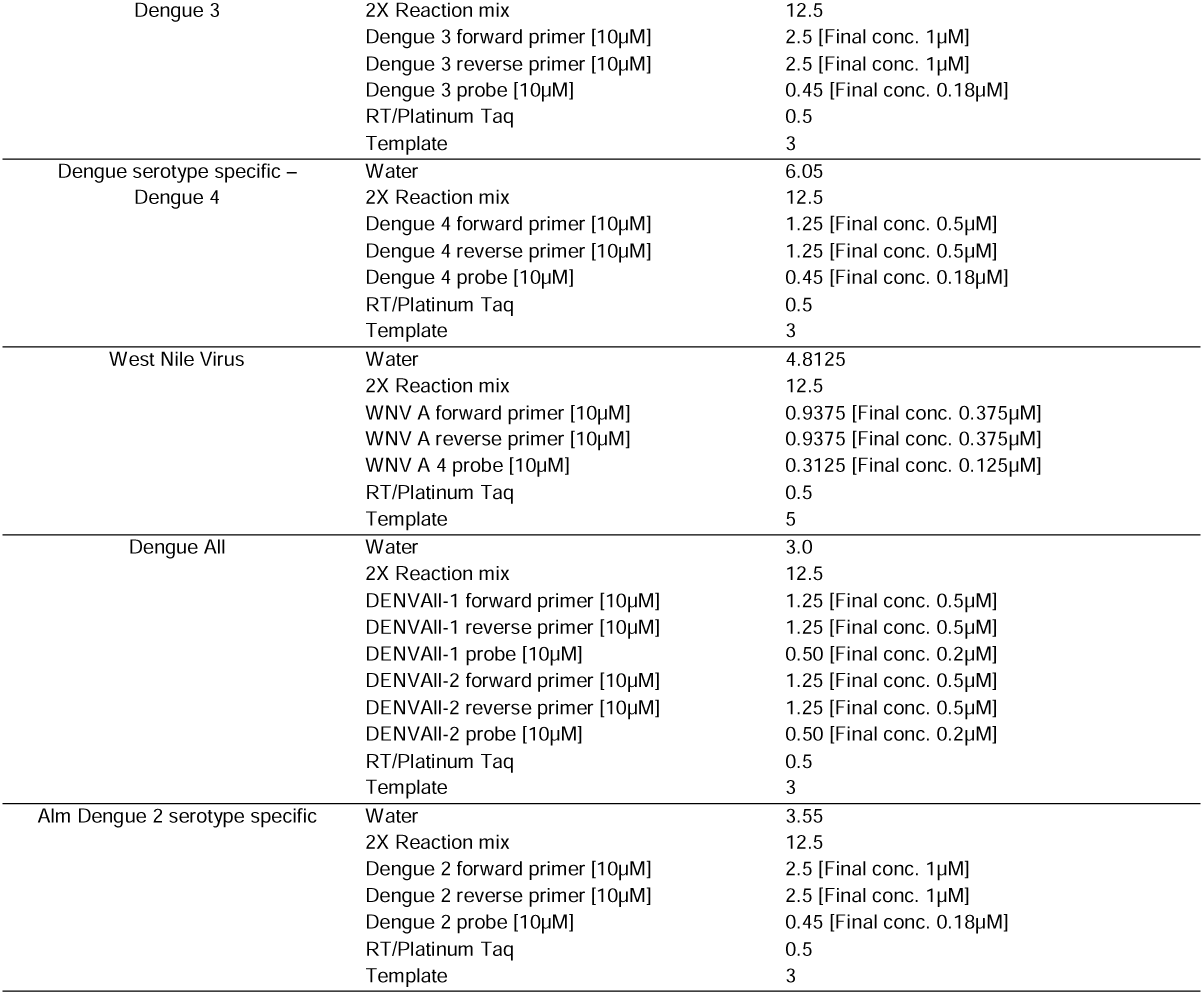
Reaction volumes for each assay using SuperScript™ III Platinum™ One-Step qRT-PCR Kit. Volumes are representative of one 25µL reaction. Primer and probes were diluted to working solutions of 10µM for assay use. Final primer and probe concentrations/reaction are indicated [ ]

**Appendix Table 5.**
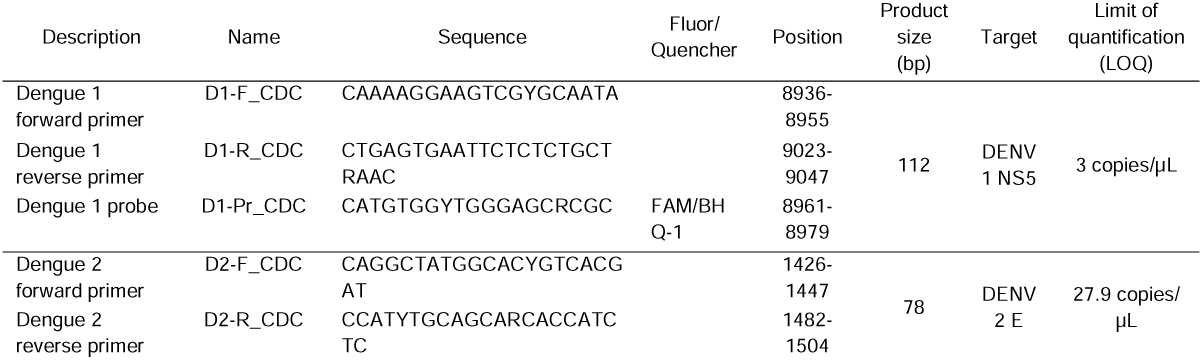
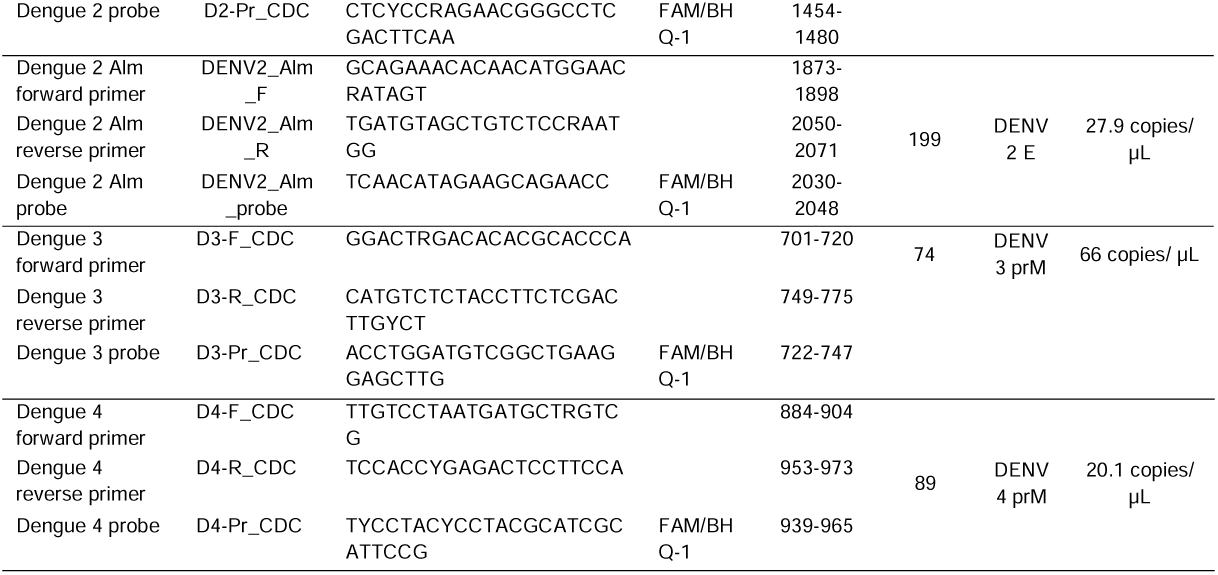
Serotype-specific assays, including both CDC (Santiago et al., 2013, doi:10.1371/journal.pntd.0002311) and Alm dengue 2 serotype-specific assays (Alm et al., 2015, doi: 10.1186/s12879-015-1226-z). “Alm” is used to refer to the publication describing the assay primers and probe to differentiate it from the CDC DENV-2 target

**Appendix Table 6.**
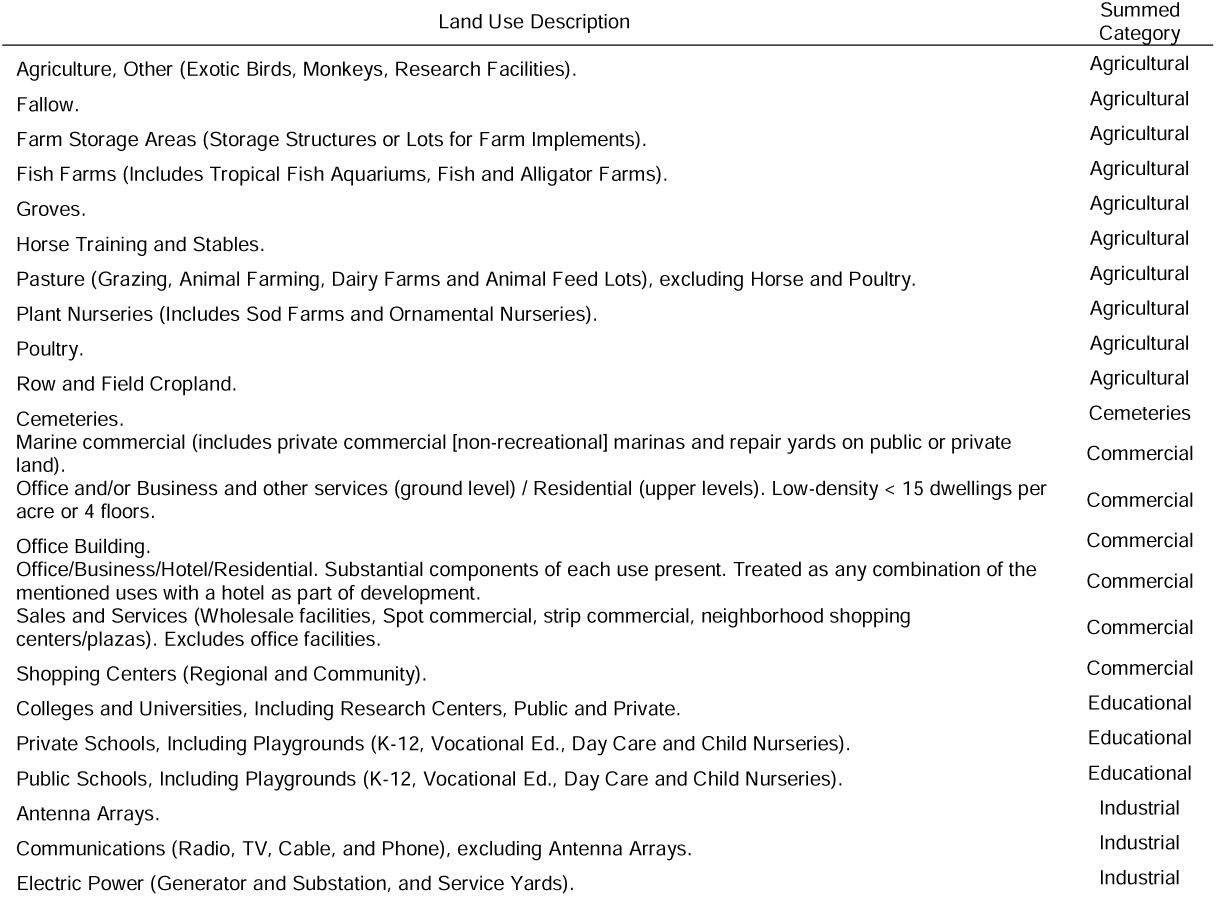
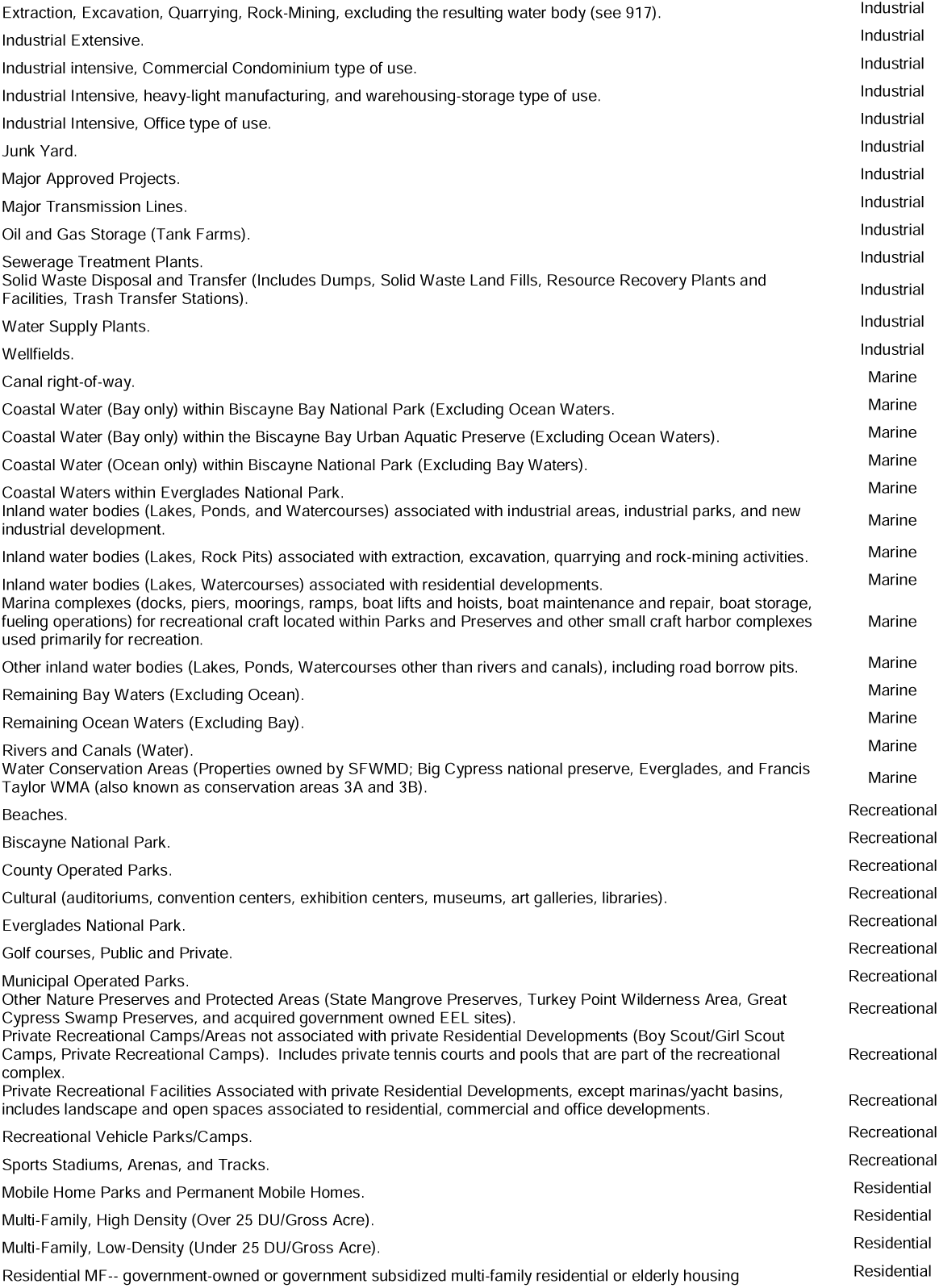
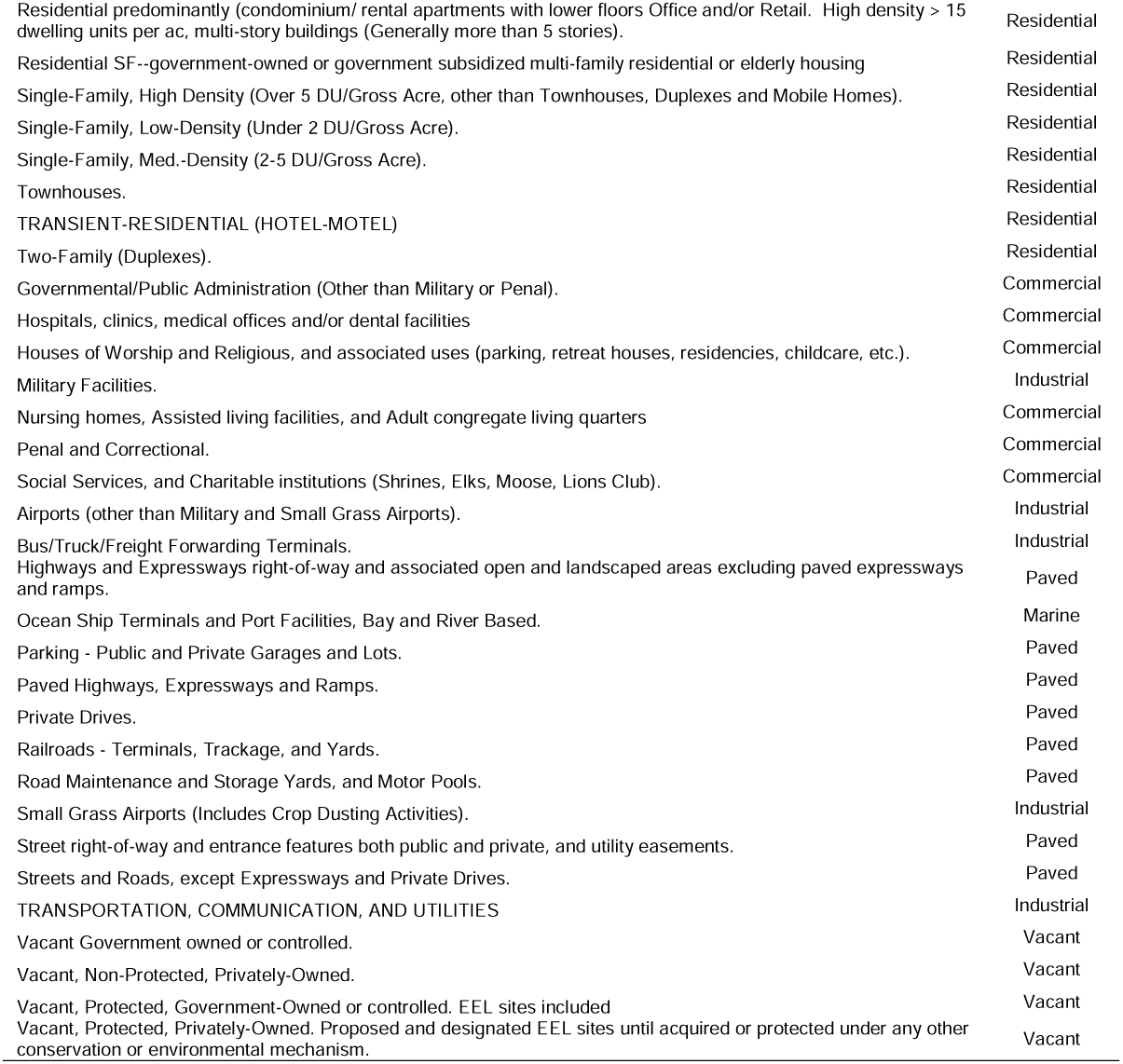
Summed categories of land use description types from the 2010 US Census

| Land Use Description | Summed<br>Category |
| --- | --- |
| Agriculture, Other (Exotic Birds, Monkeys, Research Facilities). | Agricultural |
| Fallow. | Agricultural |
| Farm Storage Areas (Storage Structures or Lots for Farm Implements). | Agricultural |
| Fish Farms (Includes Tropical Fish Aquariums, Fish and Alligator Farms). | Agricultural |
| Groves. | Agricultural |
| Horse Training and Stables. | Agricultural |
| Pasture (Grazing, Animal Farming, Dairy Farms and Animal Feed Lots), excluding Horse and Poultry. | Agricultural |
| Plant Nurseries (Includes Sod Farms and Ornamental Nurseries). | Agricultural |
| Poultry. | Agricultural |
| Row and Field Cropland. | Agricultural |
| Cemeteries. | Cemeteries |
| Marine commercial (includes private commercial [non-recreational] marinas and repair yards on public or private land). | Commercial |
| Office and/or Business and other services (ground level) / Residential (upper levels). Low-density < 15 dwellings per acre or 4 floors. | Commercial |
| Office Building. | Commercial |
| Office/Business/Hotel/Residential. Substantial components of each use present. Treated as any combination of the mentioned uses with a hotel as part of development. | Commercial |
| Sales and Services (Wholesale facilities, Spot commercial, strip commercial, neighborhood shopping centers/plazas). Excludes office facilities. | Commercial |
| Shopping Centers (Regional and Community). | Commercial |
| Colleges and Universities, Including Research Centers, Public and Private. | Educational |
| Private Schools, Including Playgrounds (K-12, Vocational Ed., Day Care and Child Nurseries). | Educational |
| Public Schools, Including Playgrounds (K-12, Vocational Ed., Day Care and Child Nurseries). | Educational |
| Antenna Arrays. | Industrial |
| Communications (Radio, TV, Cable, and Phone), excluding Antenna Arrays. | Industrial |
| Electric Power (Generator and Substation, and Service Yards). | Industrial |
| Extraction, Excavation, Quarrying, Rock-Mining, excluding the resulting water body (see 917). | Industrial |
| Industrial Extensive. | Industrial |
| Industrial intensive, Commercial Condominium type of use. | Industrial |
| Industrial Intensive, heavy-light manufacturing, and warehousing-storage type of use. | Industrial |
| Industrial Intensive, Office type of use. | Industrial |
| Junk Yard. | Industrial |
| Major Approved Projects. | Industrial |
| Major Transmission Lines. | Industrial |
| Oil and Gas Storage (Tank Farms). | Industrial |
| Sewerage Treatment Plants. | Industrial |
| Solid Waste Disposal and Transfer (Includes Dumps, Solid Waste Land Fills, Resource Recovery Plants and Facilities, Trash Transfer Stations). | Industrial |
| Water Supply Plants. | Industrial |
| Wellfields. | Industrial |
| Canal right-of-way. | Marine |
| Coastal Water (Bay only) within Biscayne Bay National Park (Excluding Ocean Waters). | Marine |
| Coastal Water (Bay only) within the Biscayne Bay Urban Aquatic Preserve (Excluding Ocean Waters). | Marine |
| Coastal Water (Ocean only) within Biscayne National Park (Excluding Bay Waters). | Marine |
| Coastal Waters within Everglades National Park. | Marine |
| Inland water bodies (Lakes, Ponds, and Watercourses) associated with industrial areas, industrial parks, and new industrial development. | Marine |
| Inland water bodies (Lakes, Rock Pits) associated with extraction, excavation, quarrying and rock-mining activities. | Marine |
| Inland water bodies (Lakes, Watercourses) associated with residential developments. | Marine |
| Marina complexes (docks, piers, moorings, ramps, boat lifts and hoists, boat maintenance and repair, boat storage, fueling operations) for recreational craft located within Parks and Preserves and other small craft harbor complexes used primarily for recreation. | Marine |
| Other inland water bodies (Lakes, Ponds, Watercourses other than rivers and canals), including road borrow pits. | Marine |
| Remaining Bay Waters (Excluding Ocean). | Marine |
| Remaining Ocean Waters (Excluding Bay). | Marine |
| Rivers and Canals (Water). | Marine |
| Water Conservation Areas (Properties owned by SFWMD; Big Cypress national preserve, Everglades, and Francis Taylor WMA (also known as conservation areas 3A and 3B). | Marine |
| Beaches. | Recreational |
| Biscayne National Park. | Recreational |
| County Operated Parks. | Recreational |
| Cultural (auditoriums, convention centers, exhibition centers, museums, art galleries, libraries). | Recreational |
| Everglades National Park. | Recreational |
| Golf courses, Public and Private. | Recreational |
| Municipal Operated Parks. | Recreational |
| Other Nature Preserves and Protected Areas (State Mangrove Preserves, Turkey Point Wilderness Area, Great Cypress Swamp Preserves, and acquired government owned EEL sites). | Recreational |
| Private Recreational Camps/Areas not associated with private Residential Developments (Boy Scout/Girl Scout Camps, Private Recreational Camps). Includes private tennis courts and pools that are part of the recreational complex. | Recreational |
| Private Recreational Facilities Associated with private Residential Developments, except marinas/yacht basins, includes landscape and open spaces associated to residential, commercial and office developments. | Recreational |
| Recreational Vehicle Parks/Camps. | Recreational |
| Sports Stadiums, Arenas, and Tracks. | Recreational |
| Mobile Home Parks and Permanent Mobile Homes. | Residential |
| Multi-Family, High Density (Over 25 DU/Gross Acre). | Residential |
| Multi-Family, Low-Density (Under 25 DU/Gross Acre). | Residential |
| Residential MF-- government-owned or government subsidized multi-family residential or elderly housing | Residential |
| Residential predominantly (condominium/ rental apartments with lower floors Office and/or Retail. High density > 15 dwelling units per ac, multi-story buildings (Generally more than 5 stories). | Residential |
| Residential SF--government-owned or government subsidized multi-family residential or elderly housing | Residential |
| Single-Family, High Density (Over 5 DU/Gross Acre, other than Townhouses, Duplexes and Mobile Homes). | Residential |
| Single-Family, Low-Density (Under 2 DU/Gross Acre). | Residential |
| Single-Family, Med.-Density (2-5 DU/Gross Acre). | Residential |
| Townhouses. | Residential |
| TRANSIENT-RESIDENTIAL (HOTEL-MOTEL) | Residential |
| Two-Family (Duplexes). | Residential |
| Governmental/Public Administration (Other than Military or Penal). | Commercial |
| Hospitals, clinics, medical offices and/or dental facilities | Commercial |
| Houses of Worship and Religious, and associated uses (parking, retreat houses, residencies, childcare, etc.). | Commercial |
| Military Facilities. | Industrial |
| Nursing homes, Assisted living facilities, and Adult congregate living quarters | Commercial |
| Penal and Correctional. | Commercial |
| Social Services, and Charitable institutions (Shrines, Elks, Moose, Lions Club). | Commercial |
| Airports (other than Military and Small Grass Airports). | Industrial |
| Bus/Truck/Freight Forwarding Terminals. | Industrial |
| Highways and Expressways right-of-way and associated open and landscaped areas excluding paved expressways and ramps. | Paved |
| Ocean Ship Terminals and Port Facilities, Bay and River Based. | Marine |
| Parking - Public and Private Garages and Lots. | Paved |
| Paved Highways, Expressways and Ramps. | Paved |
| Private Drives. | Paved |
| Railroads - Terminals, Trackage, and Yards. | Paved |
| Road Maintenance and Storage Yards, and Motor Pools. | Paved |
| Small Grass Airports (Includes Crop Dusting Activities). | Industrial |
| Street right-of-way and entrance features both public and private, and utility easements. | Paved |
| Streets and Roads, except Expressways and Private Drives. | Paved |
| TRANSPORTATION, COMMUNICATION, AND UTILITIES | Industrial |
| Vacant Government owned or controlled. | Vacant |
| Vacant, Non-Protected, Privately-Owned. | Vacant |
| Vacant, Protected, Government-Owned or controlled. EEL sites included | Vacant |
| Vacant, Protected, Privately-Owned. Proposed and designated EEL sites until acquired or protected under any other conservation or environmental mechanism. | Vacant |

**Appendix Table 7.**
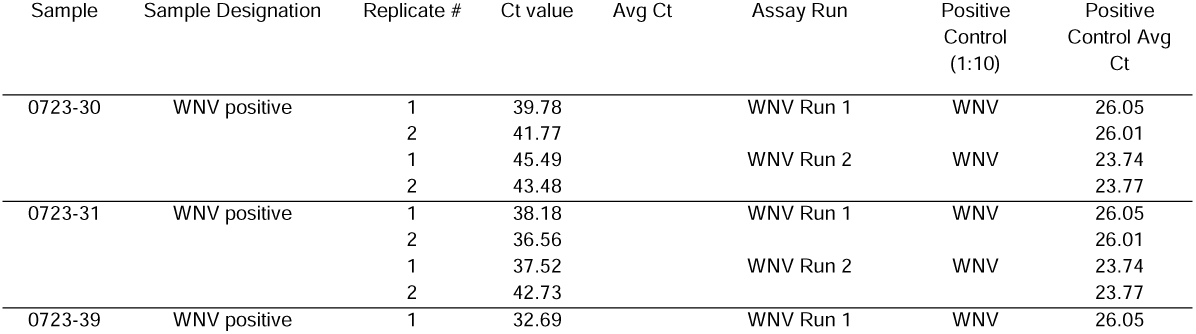
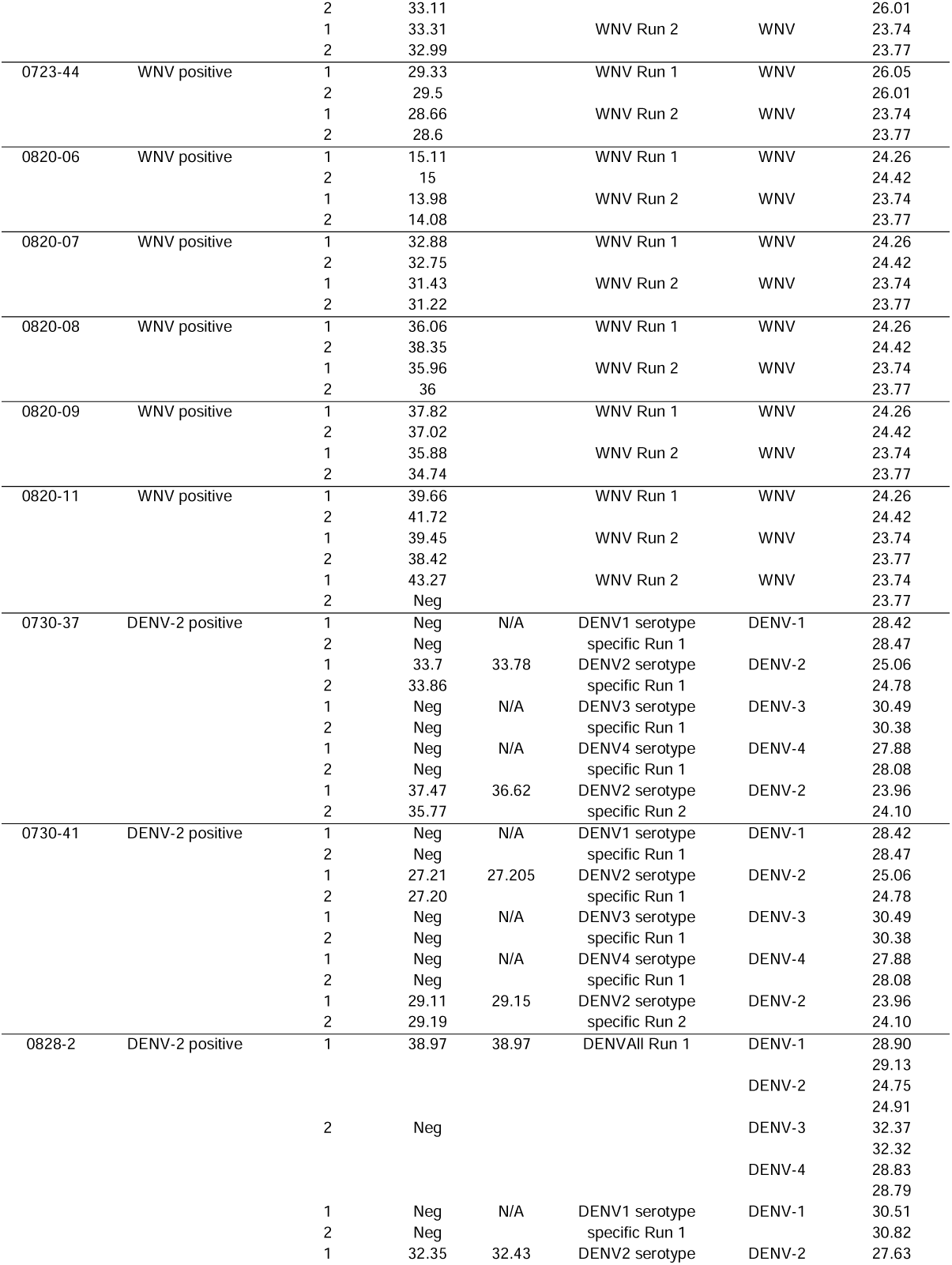
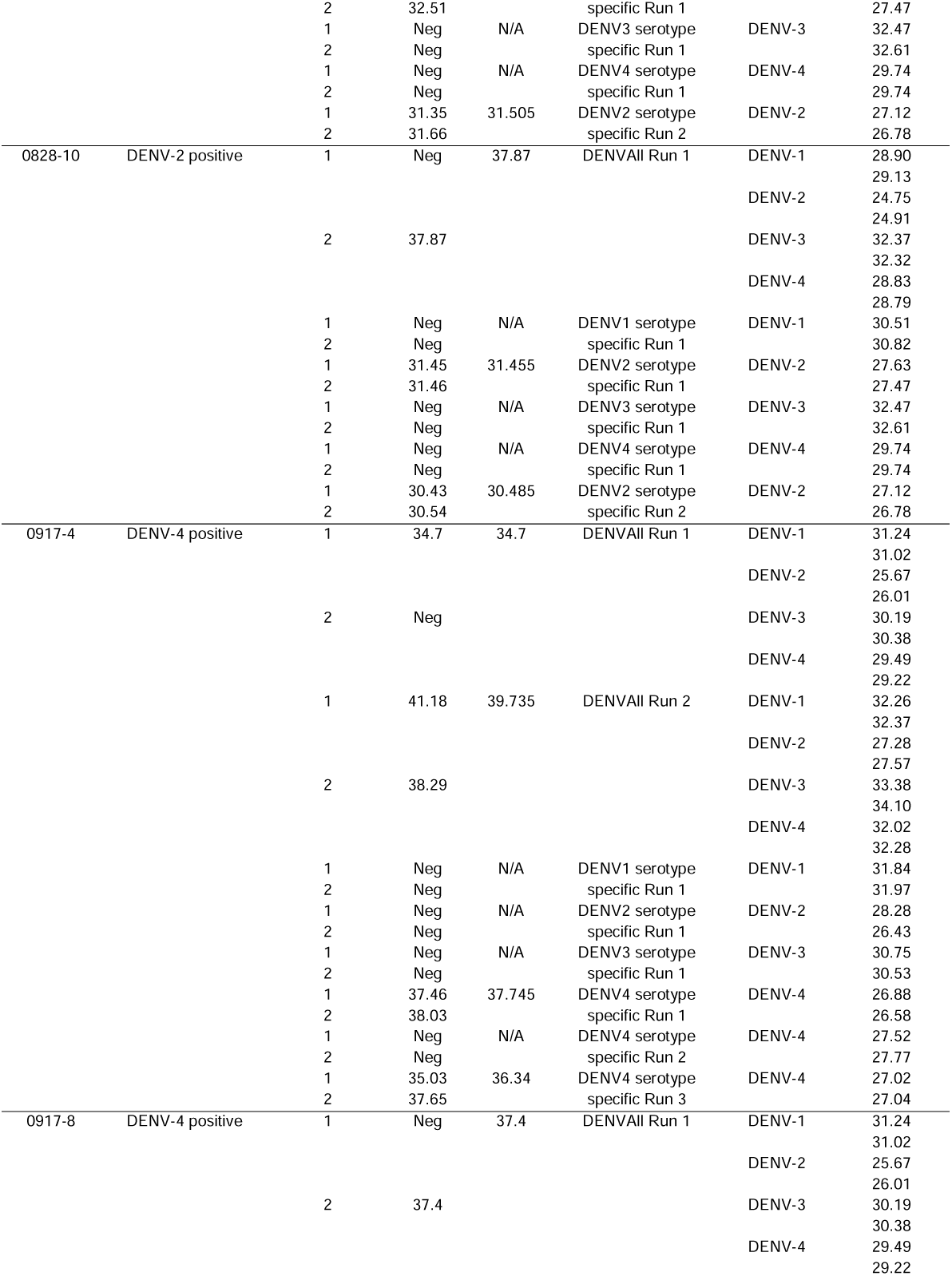
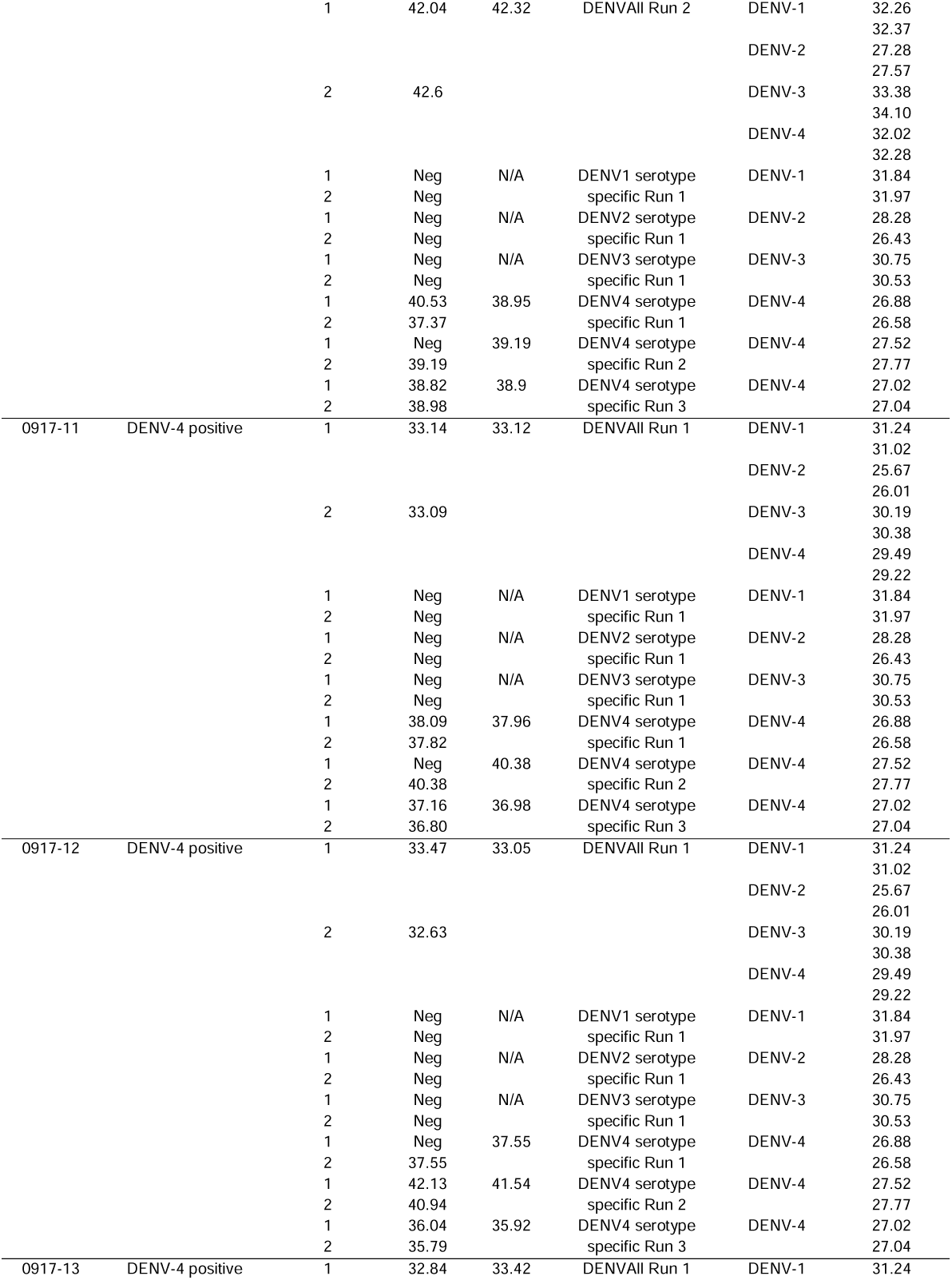
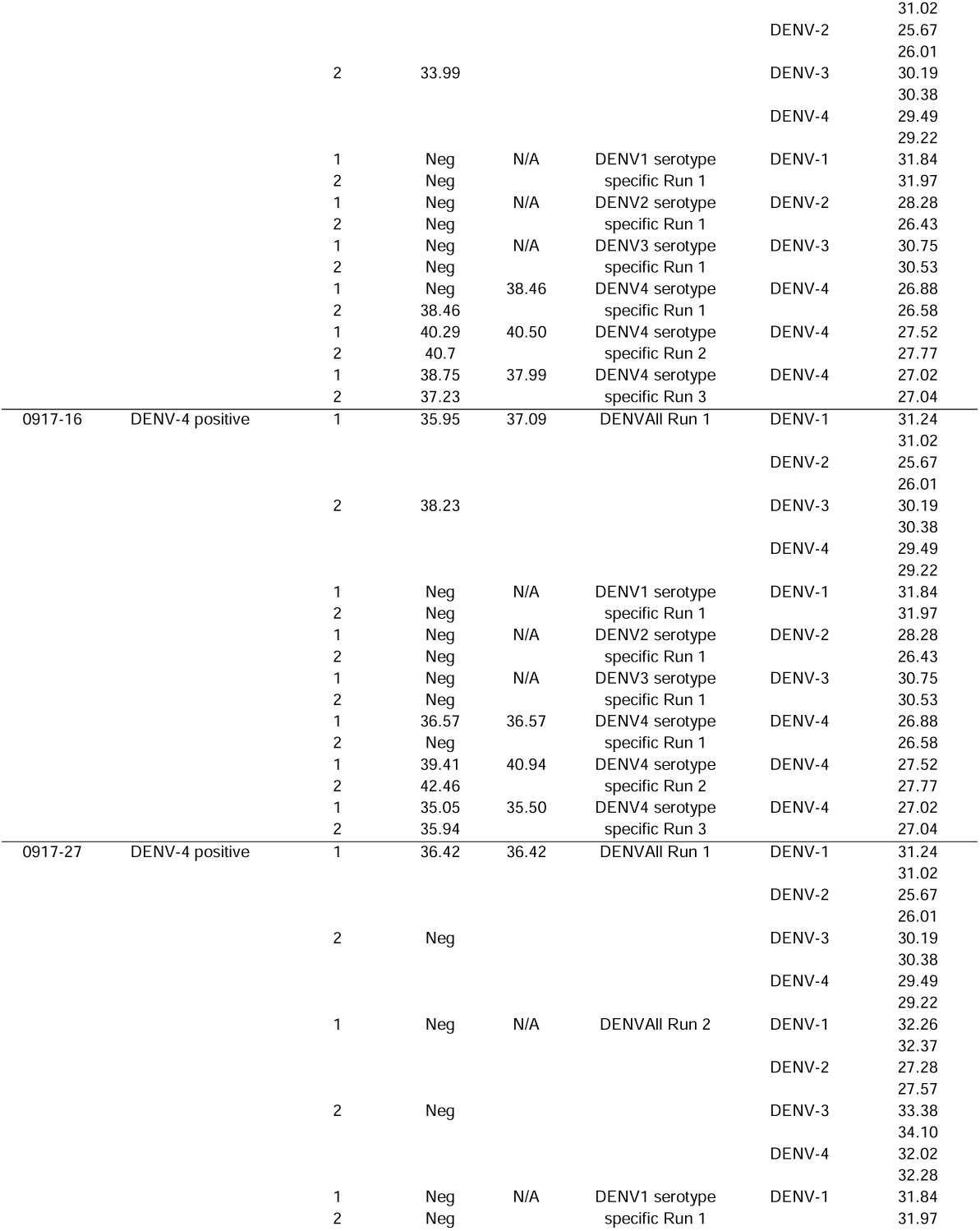
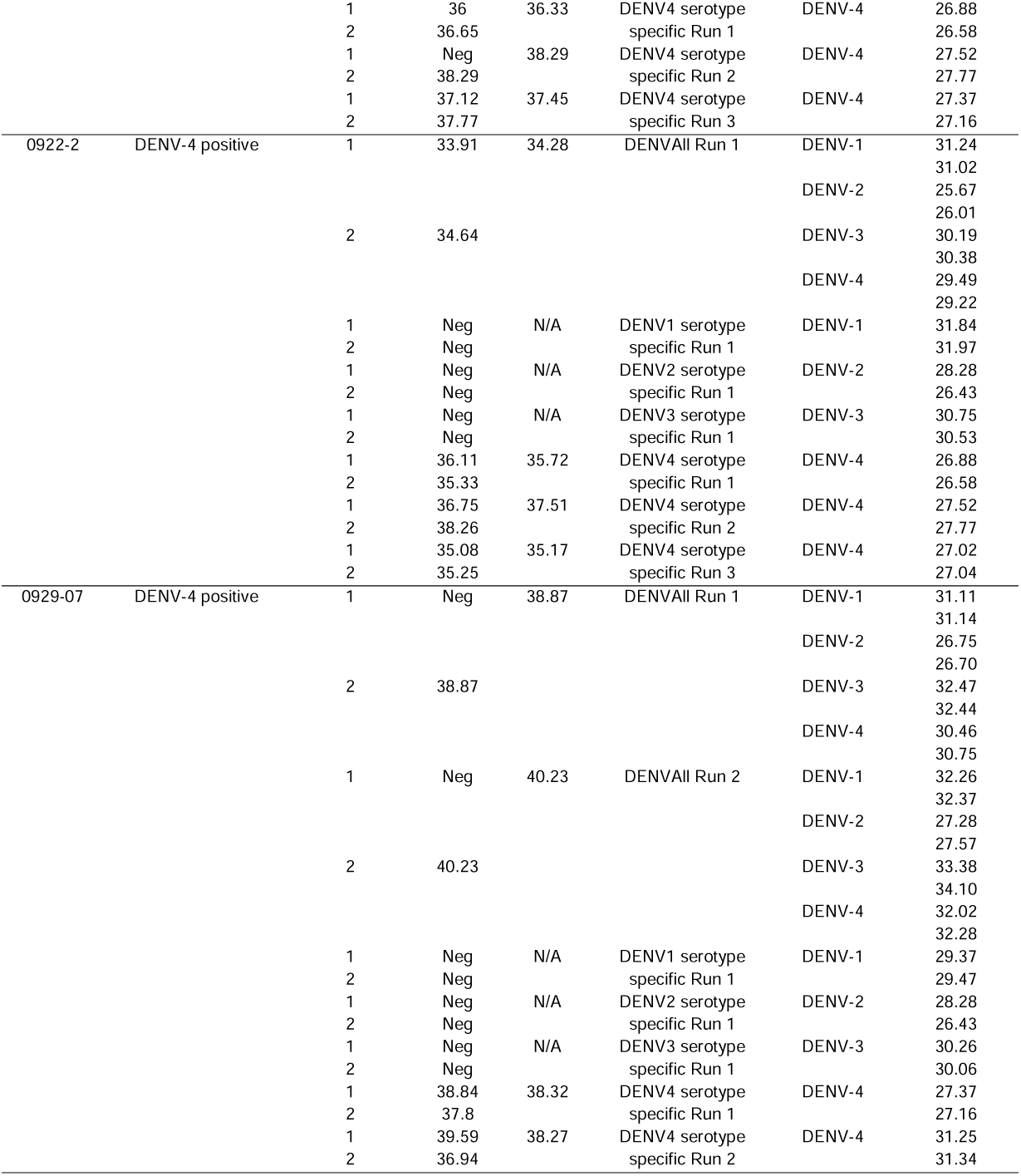
WNV-positive and DENV-positive samples

**Appendix Figure 1.**
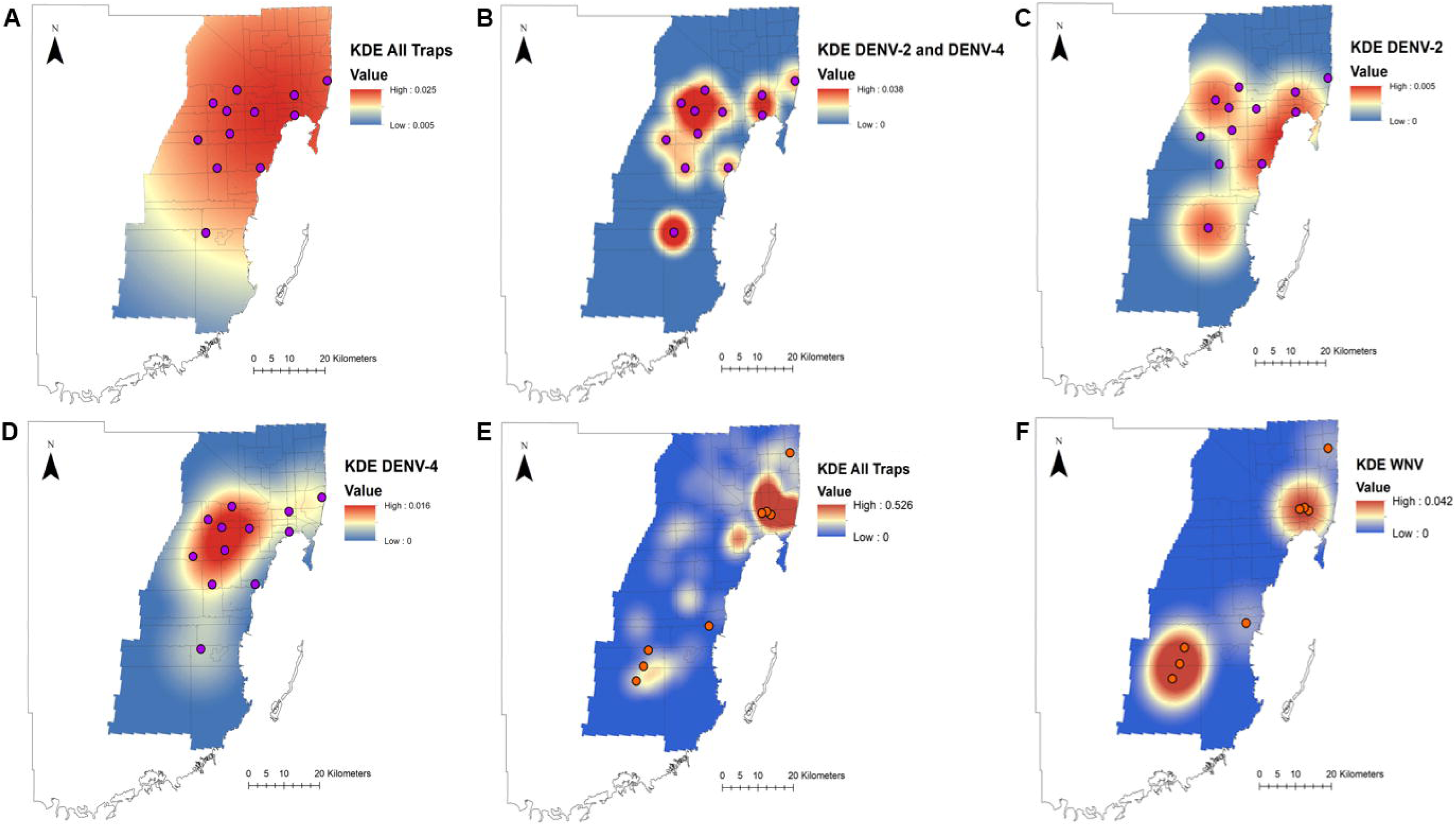
Kernel density estimation (KDE) of uninfected vector populations and arbovirus positive mosquito pools (purple – DENV, orange – WNV). A: KDE on traps that collected DENV vectors, B: KDE on all DENV positive pools, C: KDE on all DENV-2 positive pools, D: KDE on all DENV-4 positive pools, E: KDE on traps that collected WNV vectors, F: KDE on WNV positive pools.

**Appendix Figure 2.**
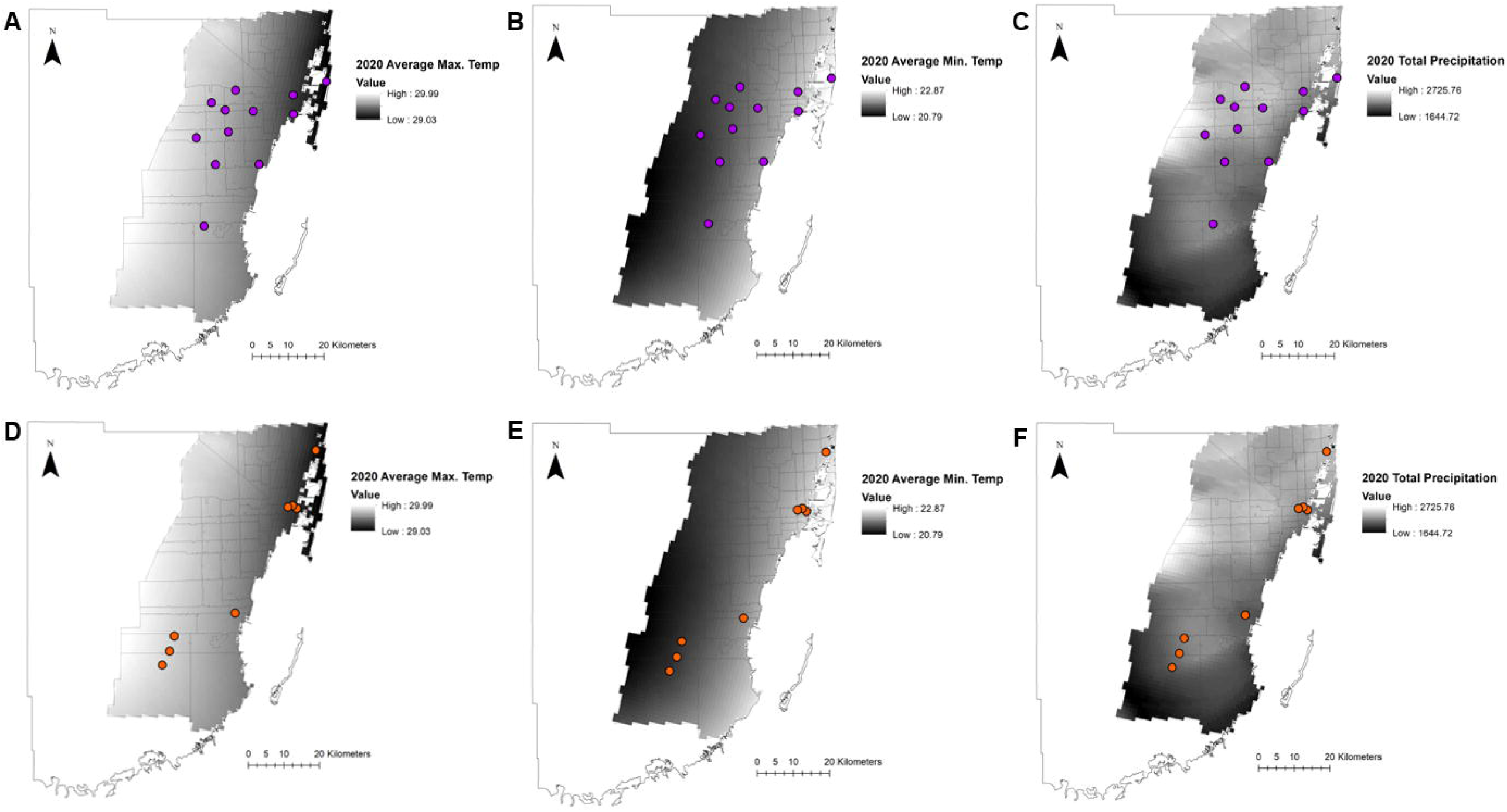
Yearly temperature and precipitation maps for Miami-Dade County in 2020 overlayed with arbovirus positive mosquito pools (purple – DENV, orange – WNV). A and D: Maximum average temperature for 2020 in Miami-Dade visualized with DENV positive pools (A) or WNV positive pools (D). B and E: Minimum average temperature for 2020 in Miami-Dade visualized with DENV positive pools (B) or WNV positive pools (E). C and F: Total precipitation in Miami-Dade in 2020 visualized with DENV positive pools (C) or WNV positive pools (F).

**Appendix Figure 3.**
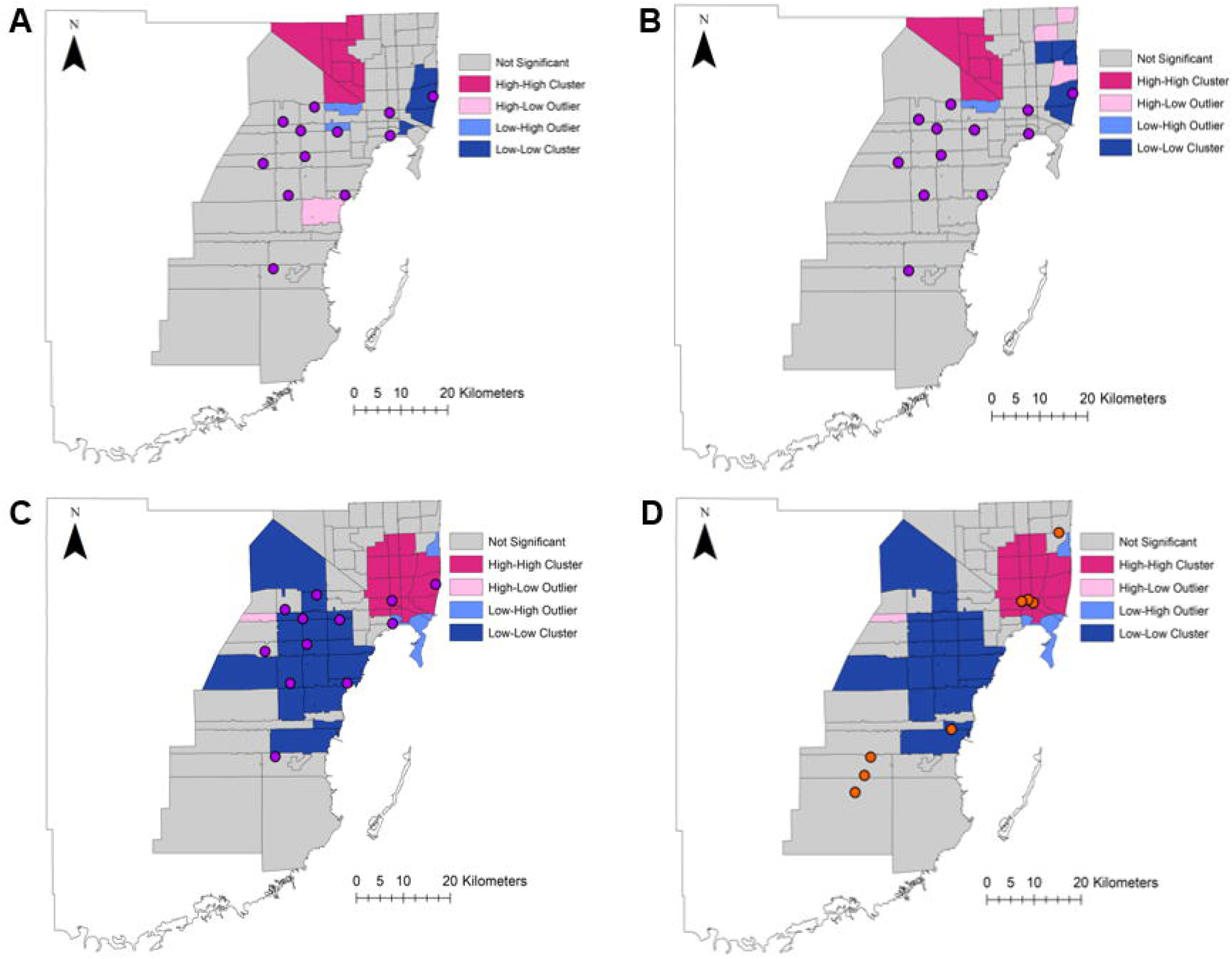
Local Moran’s I, a local indicator of spatial association (LISA), analyses of imported DENV cases and PLWH overlayed with arbovirus positive mosquito pools (purple – DENV, orange – WNV). A: LISA from DENV imported cases (2009 – 2019) visualized with 2020 DENV positive pools, B: LISA from 2019 DENV imported cases visualized with 2020 DENV positive pools, C: LISA from PLWH visualized with 2020 DENV positive pools, D: LISA from PLWH visualized with 2020 WNV positive pools.

